# Current and emerging polymyxin resistance diagnostics: a systematic review of established and novel detection methods

**DOI:** 10.1101/2020.08.23.20180133

**Authors:** Tumisho Mmatumelo Seipei Leshaba, Nontombi Marylucy Mbelle, John Osei Sekyere

**Affiliations:** Department of Medical Microbiology, School of Medicine, Faculty of Health Sciences, University of Pretoria, South Africa

**Keywords:** Colistin resistance, detection methods, *mcr*, diagnostics, Polymyxins

## Abstract

**Background:** The emergence of polymyxin resistance, due to transferable *mcr*-genes, threatens public and animal health as there are limited therapeutic options. As polymyxin is one of the last-line antibiotics, there is a need to contain the spread of its resistance to conserve its efficacy. Herein, we describe current and emerging polymyxin resistance diagnostics to inform faster clinical diagnostic choices.

**Methods:** A literature search in diverse databases for studies published between 2016 and 2020 was performed. English articles evaluating colistin resistance methods/diagnostics were included.

**Results:** Screening resulted in the inclusion of 93 journal articles. Current colistin resistance diagnostics are either phenotypic or molecular. Broth microdilution (BMD) is currently the only gold standard for determining colistin MICs (minimum inhibitory concentration).

Phenotypic methods comprise of agar-based methods such as CHROMagar™ Col-*APSE*, SuperPolymyxin, ChromID® Colistin R, LBJMR, and LB medium; manual MIC-determiners viz., UMIC, MICRONAUT MIC-Strip (MMS), and ComASP Colistin; automated antimicrobial susceptibility testing (AST) systems such as BD Phoenix, MICRONAUT-S, MicroScan, Sensititre and Vitek 2; MCR-detectors such as lateral flow immunoassay (LFI) and chelator-based assays including EDTA- and DPA-based tests *i.e*. combined disk test (CDT), modified colistin broth-disk elution (CBDE), Colispot, and Colistin MAC test as well as biochemical colorimetric tests *i.e.* Rapid Polymyxin NP test and Rapid ResaPolymyxin NP test. Molecular methods only characterize mobile colistin resistance; they include PCR, LAMP, and whole-genome sequencing (WGS).

**Conclusion:** Due to the faster turnaround time (≤3h), improved sensitivity (84-100%), and specificity (93.3-100%) of the Rapid ResaPolymyxin NP test, we recommend this test for initial screening of colistin-resistant isolates. This can be followed by CBDE with EDTA or the LFI as they both have 100% sensitivity and a specificity of ≥ 94.3% for the rapid screening of *mcr*-genes. However, molecular assays such as LAMP and PCR may be considered in well-equipped clinical laboratories.

**Author summary/highlights/importance:**

- Polymyxin resistance is rapidly increasing, threatening public and veterinary healthcare.
- As one of the last-line antibiotics, polymyxin must be conserved by containing the spread of polymyxin resistance.
- Detecting colistin resistance relies on determining colistin MIC values by standard broth microdilution, which is labour-intensive with longer turnaround time (TAT).
- Other polymyxin resistance diagnostics have been developed to augment or replace the broth microdilution with faster TAT.
- Based on their respective sensitivities, specificities, TAT, skill, and cost, selected phenotypic and molecular assays are recommended for laboratories, according to their financial strengths, to enhance colistin resistance surveillance and control.

## 1. Introduction

The rapid dissemination of multidrug-resistant (MDR) *Acinetobacter spp*., *Pseudomonas spp*. and carbapenemase-producing *Enterobacterales* (CPE) has been of vital significance to public and veterinary health ^1,2^. Of particular concern are carbapenem-resistant infections caused by these organisms ^3^, as they are associated with high mortality rates owing to limited therapeutic options ^4,5^. The limited pipeline of new antibiotic classes has led to increased use of polymyxin E (colistin) alone or in combination with tigecycline or fosfomycin for the treatment of MDR Gram-negative infections ^6, 7^.

Polymyxin (colistin) is of particular value as a last-line antibiotic for treating MDR and carbapenem-resistant Gram-negative infections as it is bactericidal unlike tigecycline, which is bacteriostatic and is not readily available in many countries ^8^. Polymyxin consists of hydrophilic and lipophilic moieties that allow it to have stable polar and hydrophobic interactions with the lipopolysaccharide (LPS) membrane of Gram-negative bacteria ^9, 10^. These interactions result in the destruction of the LPS membrane, causing the cytoplasmic content to leak out, ultimately killing the cell ^10^. There are two types of polymyxins, B and E, but this review shall focus on polymyxin E, also known as colistin.

Increased use of polymyxin to treat MDR Gram-negative infections has led to the emergence of acquired colistin resistance ^11^. Several mechanisms that mediate acquired colistin resistance have been identified, the most common being chromosomal mutations and plasmid-borne colistin resistance ^10, 12, 13^. Chromosomal mutations result in modification(s) of the LPS membrane using different mechanisms: (i) the addition of 4-amino-4-deoxy-L-arabinose (L-Ara-4N), phosphoethanolamine (pETN) or galactosamine moieties at the 4’ or 1’ position of lipid A, which reduces the overall anionic charge of the LPS; (ii) overexpression of efflux pumps systems; (iii) overproduction of capsule polysaccharide that reduces the LPS membrane’s permeability ^8, 12^.

Plasmid-borne colistin resistance involves the acquisition of a mobile colistin resistance (*mcr*) gene that encodes a pETN transferase ^7^. Since the discovery of the first plasmid-borne *mcr*-1 gene in *Escherichia coli* in China, other *mcr* variants viz., *mcr*-2, *mcr*-3, *mcr*-4, *mcr*-5, *mcr*-6, *mcr*-7, *mcr*-8, *mcr*-9, and *mcr*-10, have been described worldwide ^6, 14 -21^. Currently, the confirmation of polymyxin E resistance relies on the broth microdilution (BMD) ^22^. Although BMD is the gold standard for colistin susceptibility testing, it is not suitable for routine clinical use as it is time-consuming and associated with methodological issues ^23^. Transmissible colistin resistance makes it imperative to establish rapid and reliable methods that will efficiently detect colistin resistance ^6^. As *mcr*-containing plasmids are capable of transfer between epidemic strains of *Enterobacterales*, rapid detection of colistin resistance could manage the dissemination of colistin resistance in human and animal populations ^6, 23^.

There has been an increasing interest in discovering alternative methods of detecting resistance to colistin arising from both chromosomal mutations and plasmid-borne *mcr* genes ^1, 24^. These methods can be categorized as either phenotypic or molecular methods ^12, 23^. This review aims to summarize and analyse clinical diagnostic methods that are currently available for detecting colistin resistance.

### Evidence before this review

Methods used to detect polymyxin resistance have been reviewed ^1, 12, 23^. Bardet and Rolain (2018) narratively described methods used to detect colistin resistance, focusing mainly on their efficiency to detect all mechanisms of colistin resistance. They also analysed methods specifically used to detect plasmid-mediated colistin resistance. Osei Sekyere (2019) provided a comprehensive description of polymyxin resistance and *mcr*-detecting diagnostic methods up to 2018. The review included the composition of culture media, primers, and cycling conditions of PCR methods. Osei Sekyere (2019) summarized the sensitivities, specificities, turnaround time (TAT), skill, relative cost, essential agreement (EA), categorical agreement (CA), major error (ME) and very major error (VME) of polymyxin-resistance detection methods. Since 2018, new evaluation studies have been reported, broadening our understanding and conclusions of the best colistin resistance diagnostics/methods. Therefore, we provide a comprehensive update and an expanded review, based on broader evaluation studies, of all the current diagnostic methods designed to detect colistin resistance.

### Literature search strategy

A comprehensive literature search was performed using Pubmed, Web of Science, and ScienceDirect. Articles published in English, from January 2016 to September 2020, were retrieved and screened using the following keywords: “colistin AND resistan*”, “polymyxin AND resistan*” in permutation and combination with “detection” and “diagnostics”, in a factorial order. The search was based on articles that were evaluating methods that are currently used for the detection of colistin resistance and *mcr*-genes. Studies based on epidemiology, risk factors, surveillance, non-English language articles, other reviews, case reports, or case studies were excluded. The inclusion and exclusion methods used in this review are demonstrated in Figure 1. The following data was extracted from the included articles and summarized in Table 1: Diagnostic methods used, types and sample size (in numbers) of bacterial species used in the evaluation, sensitivity, specificity, EA, CA, ME, VME, relative cost and TAT.

**Table 1.**
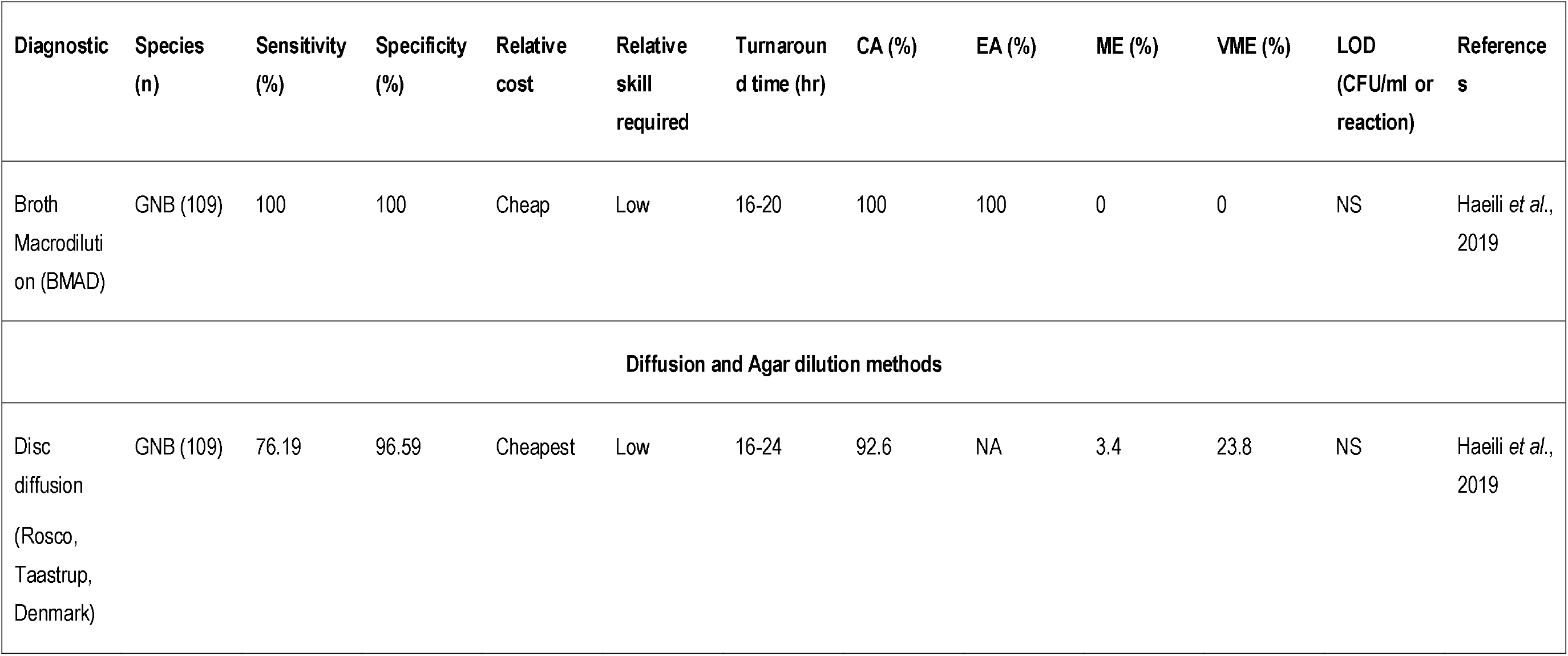
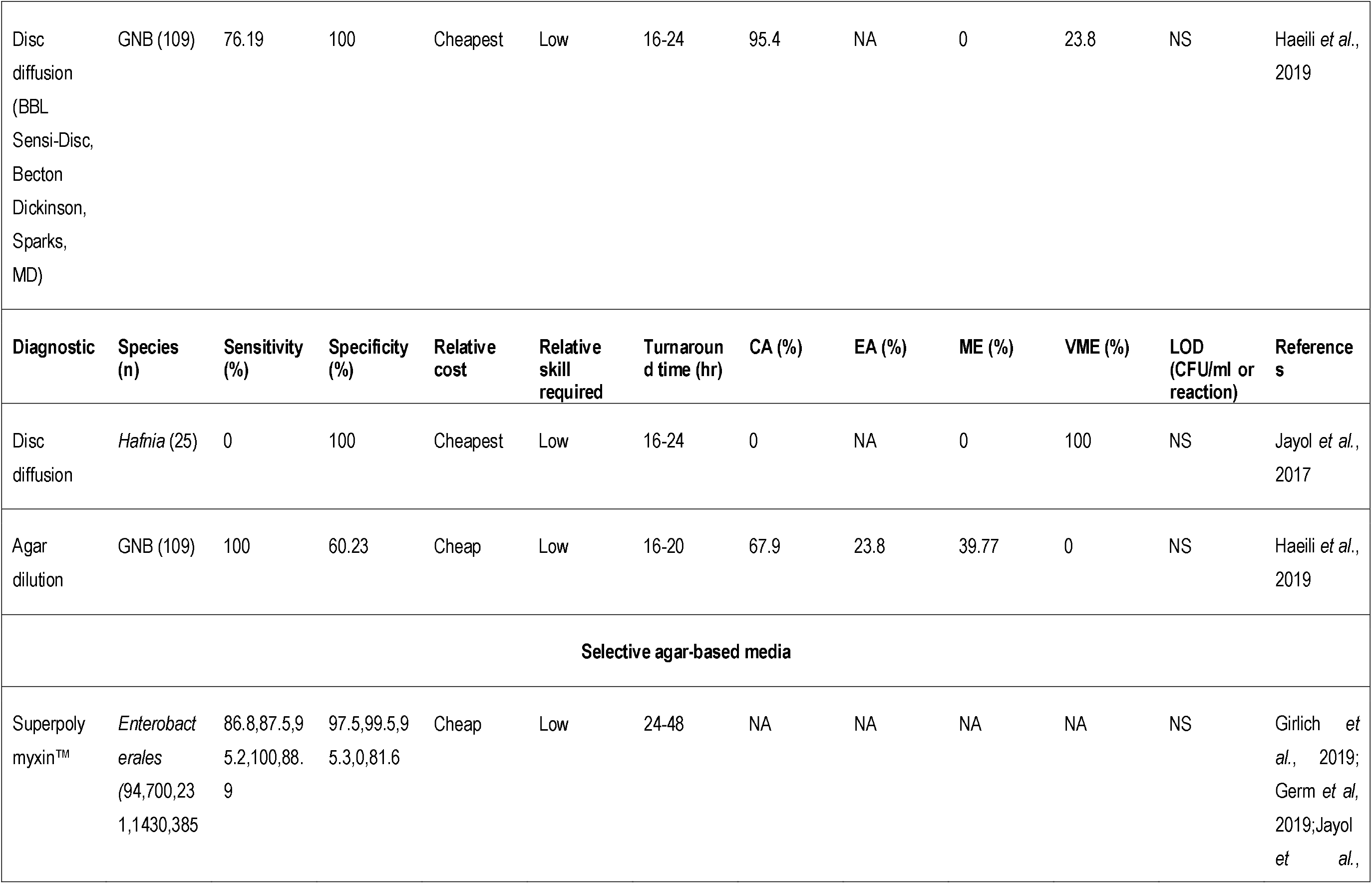
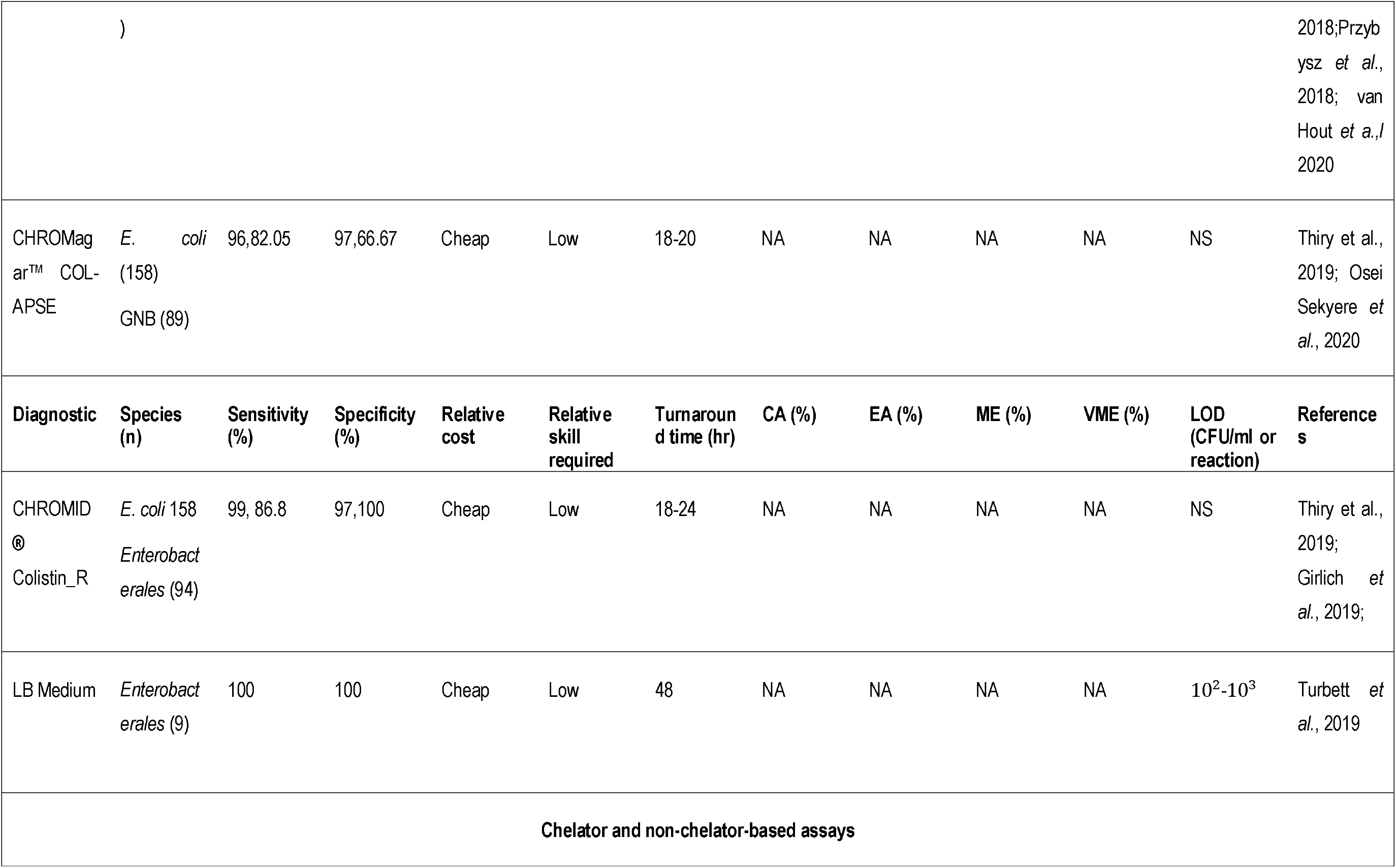
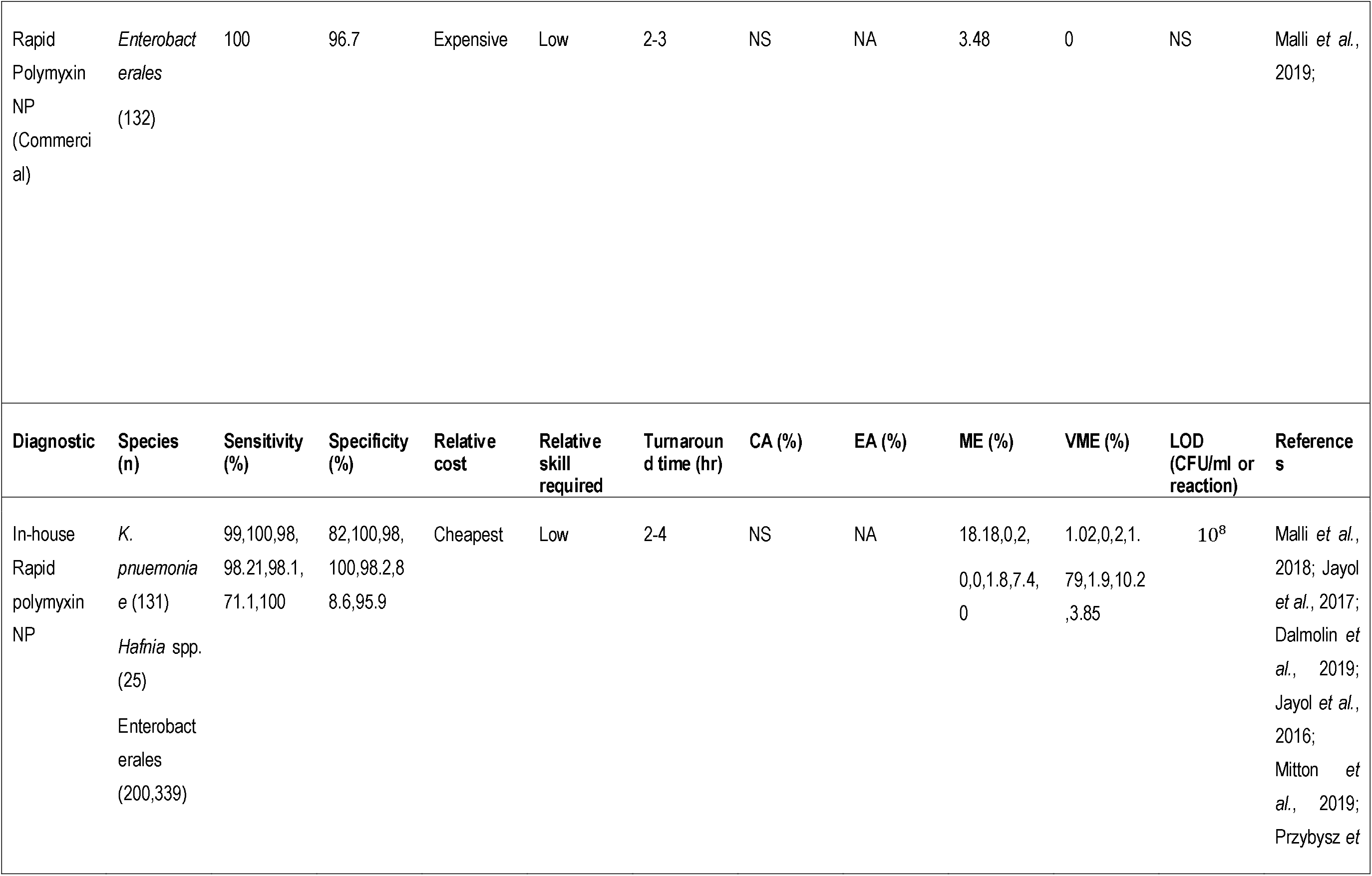
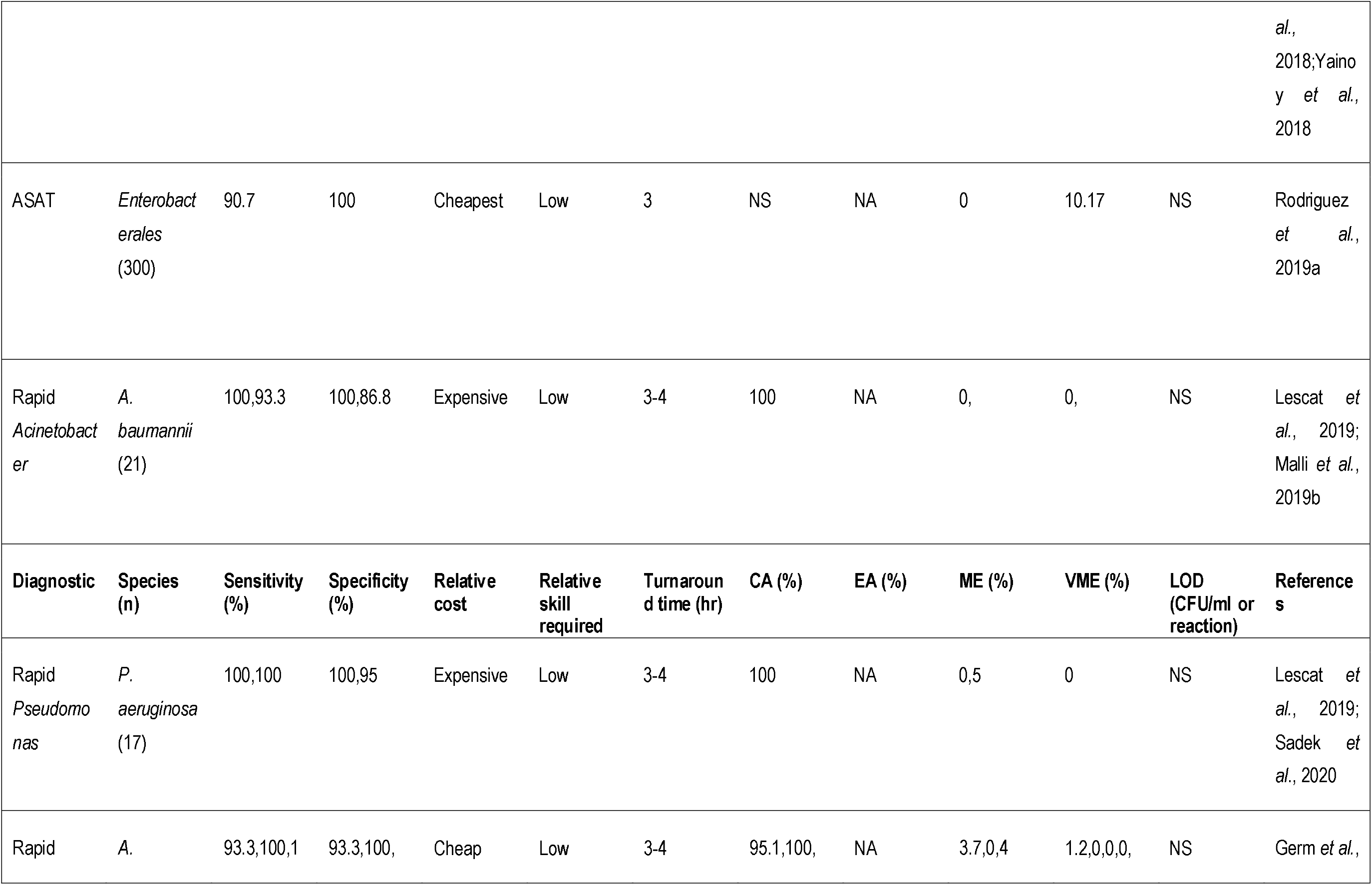
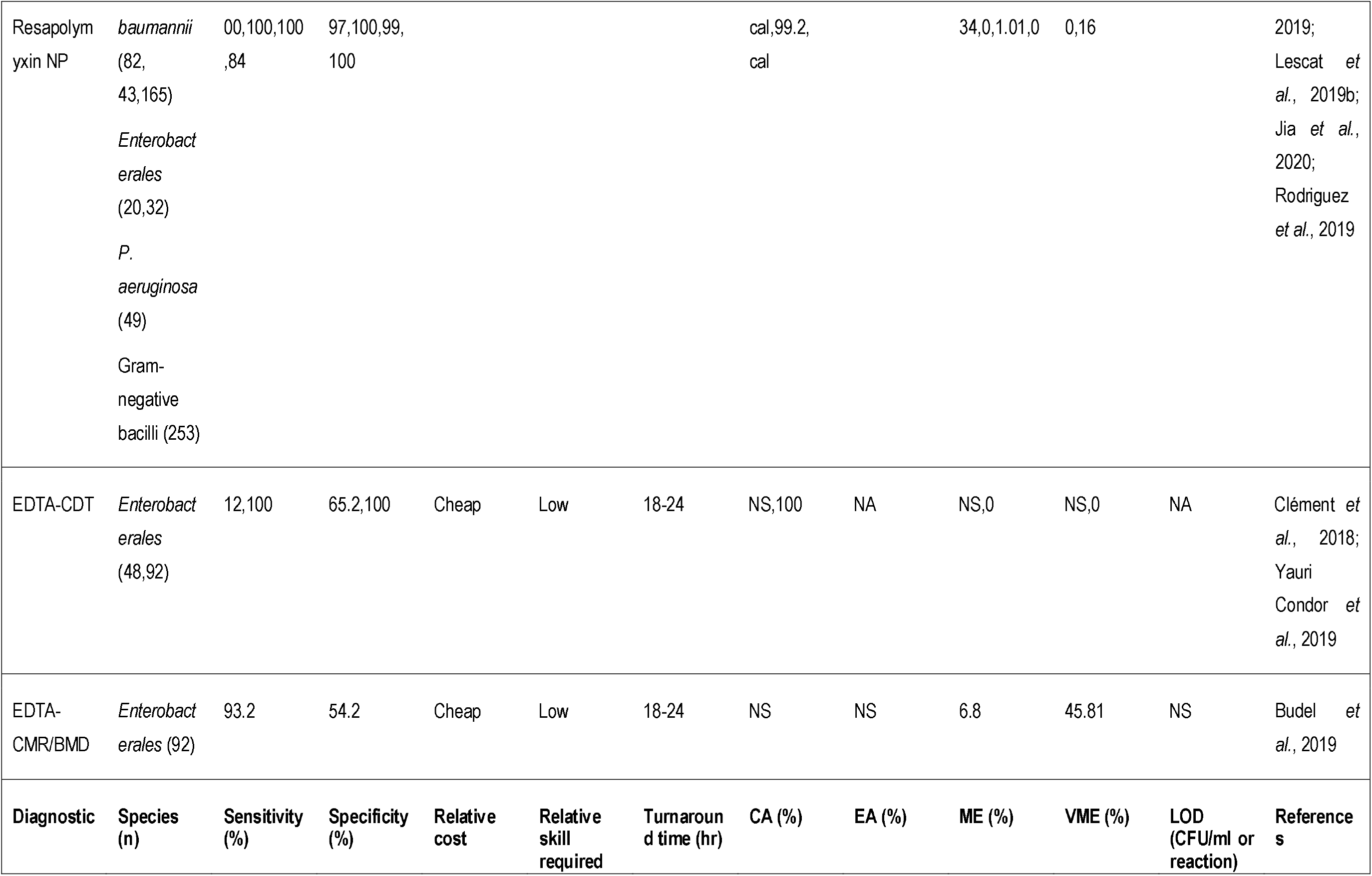
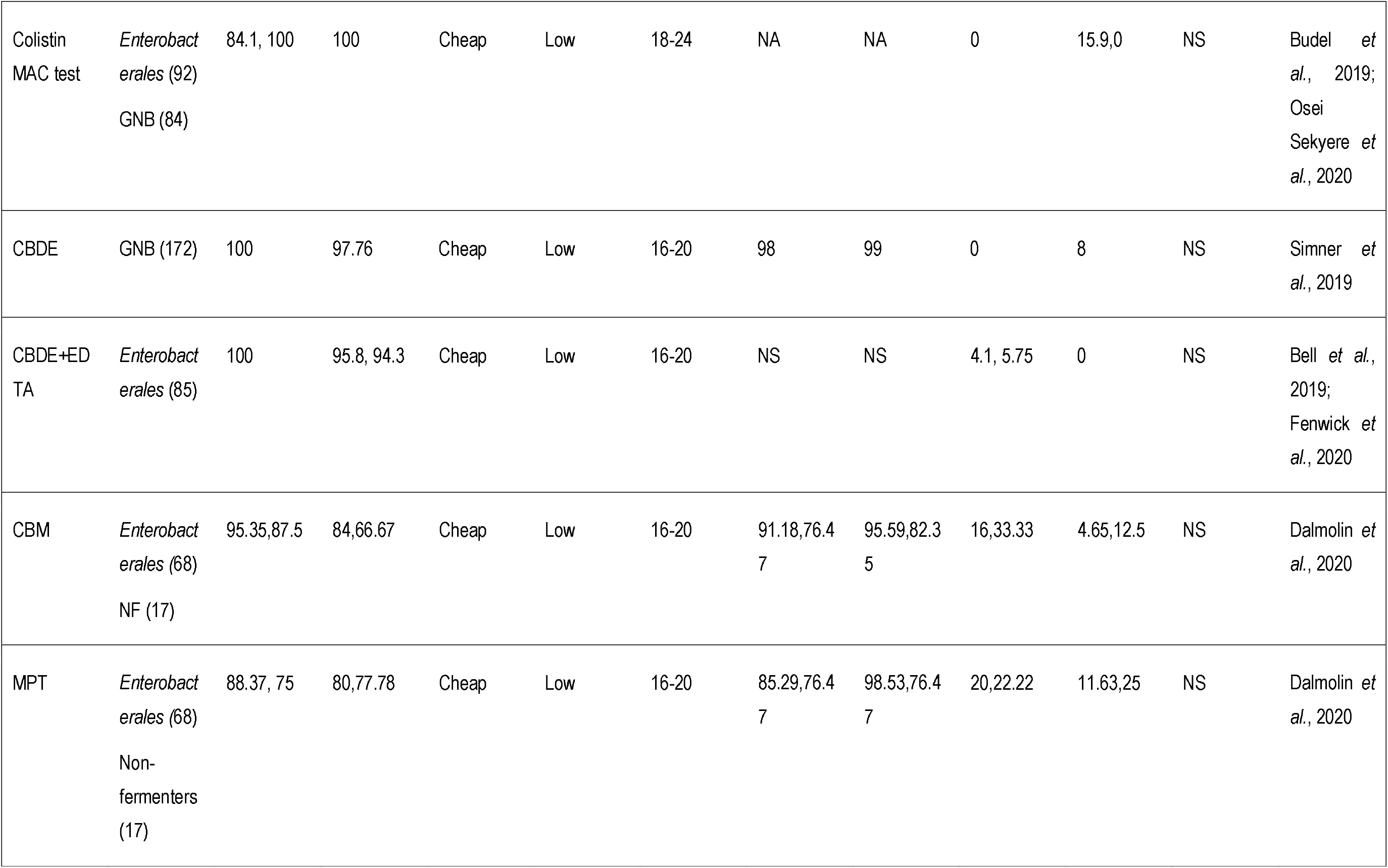
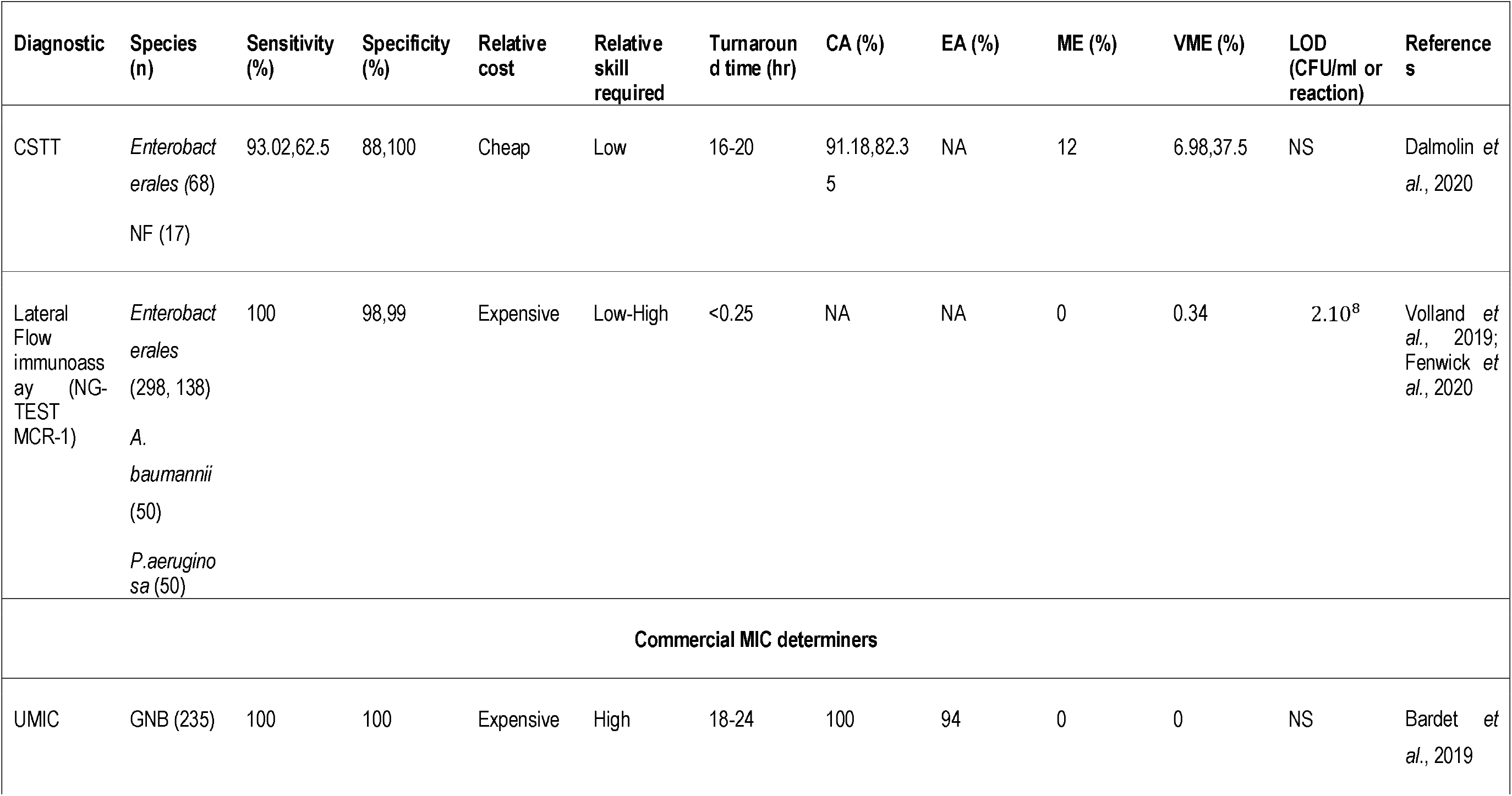
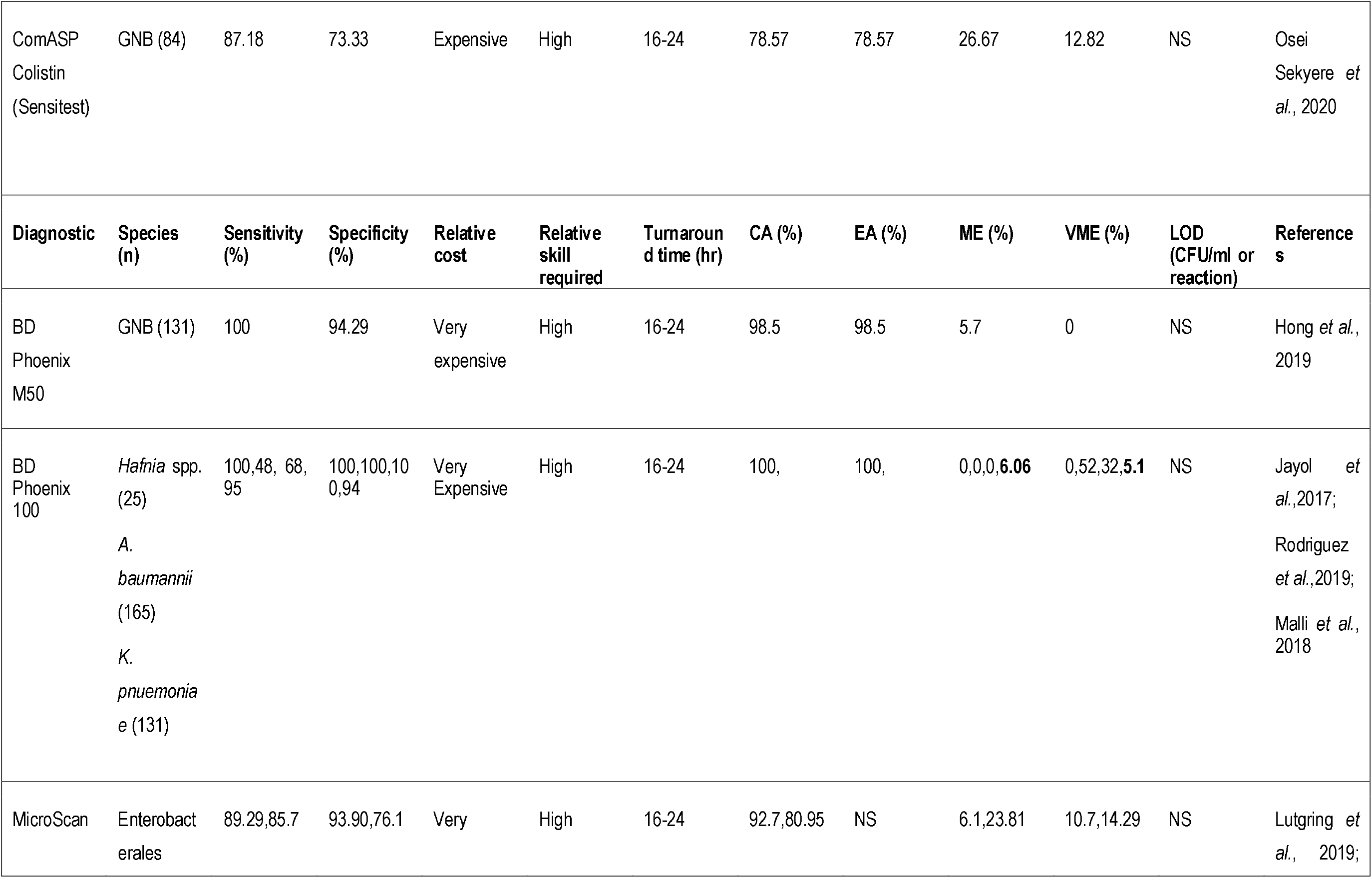
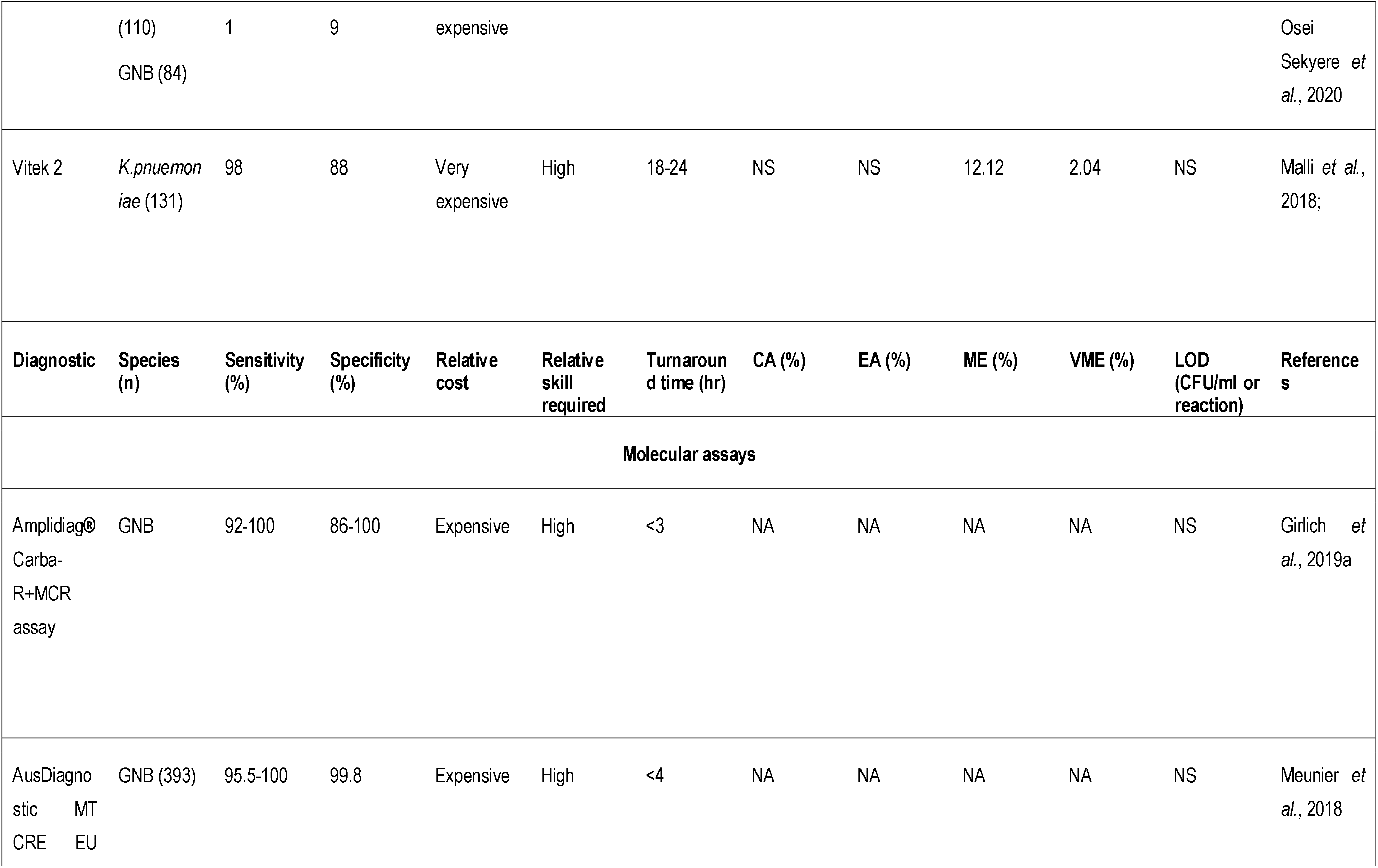
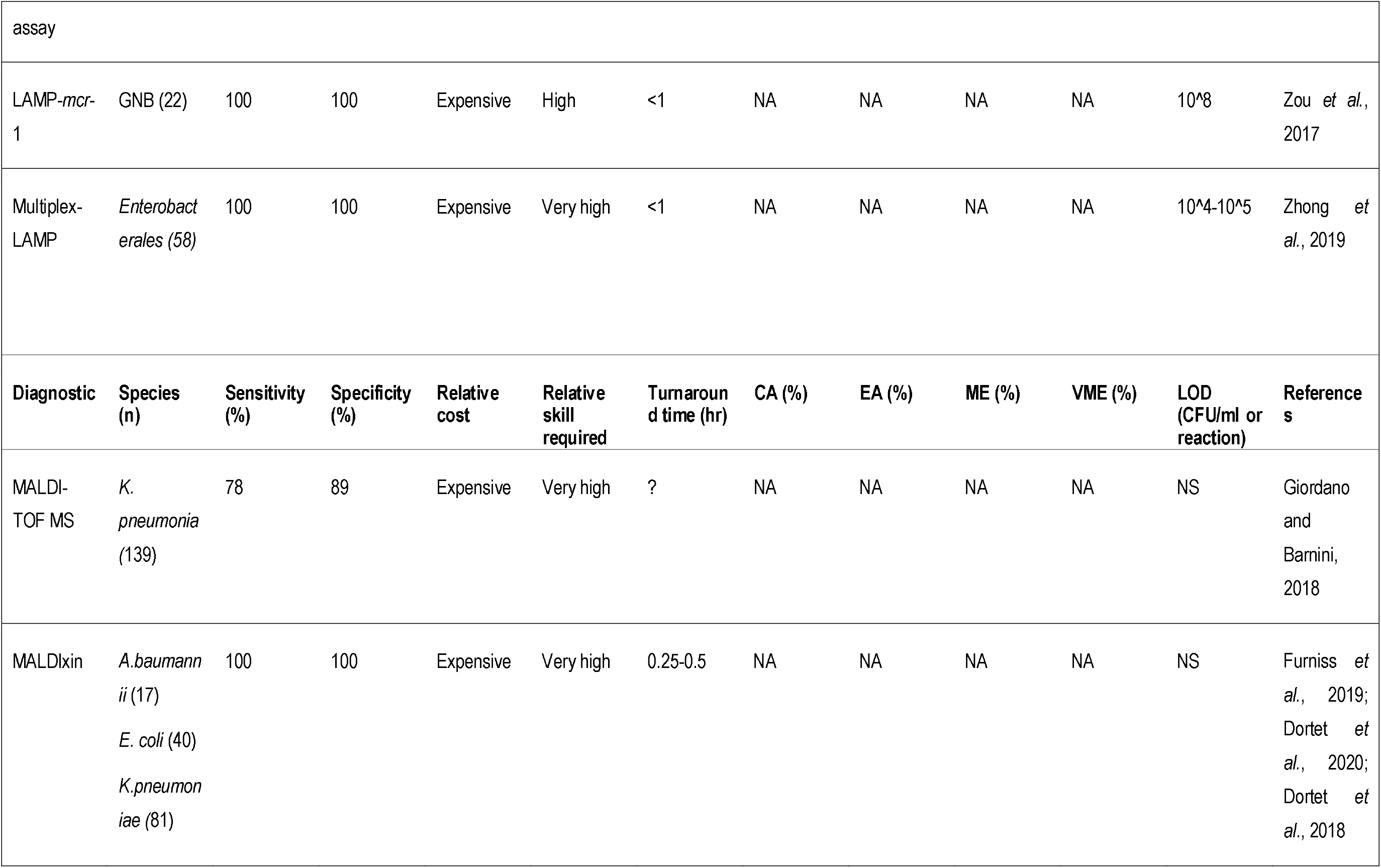
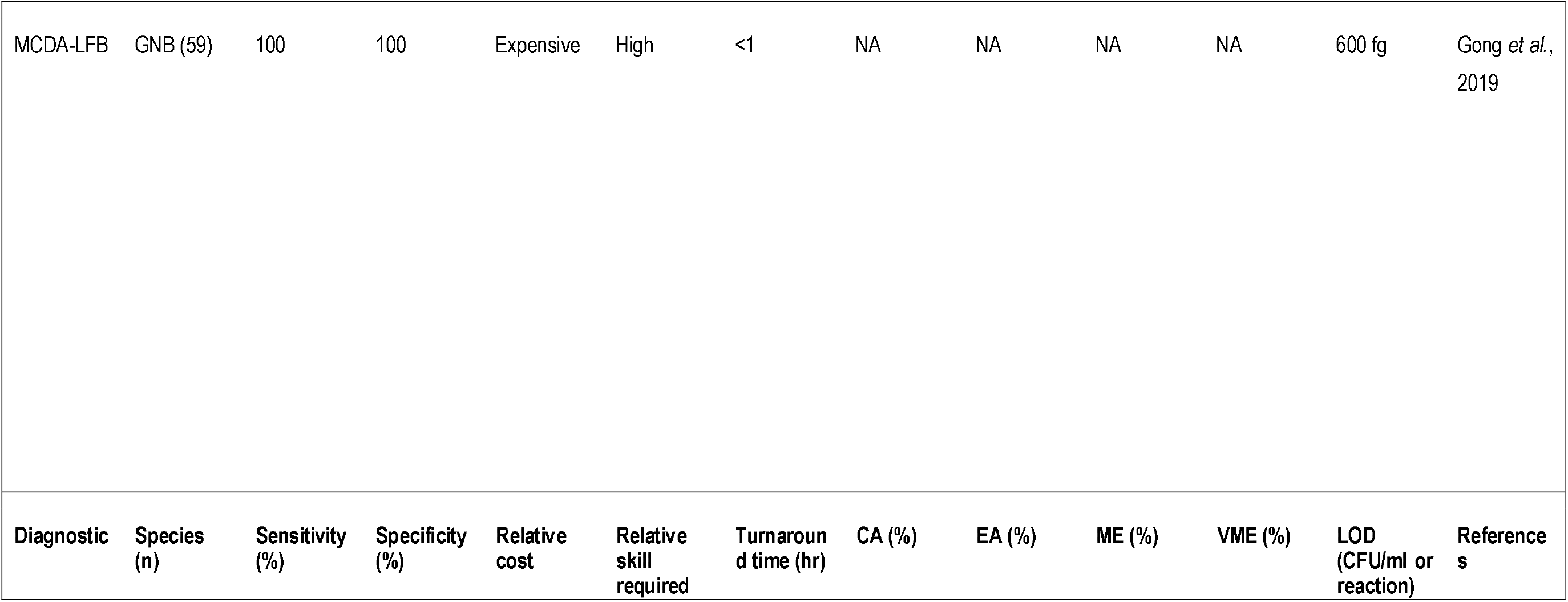
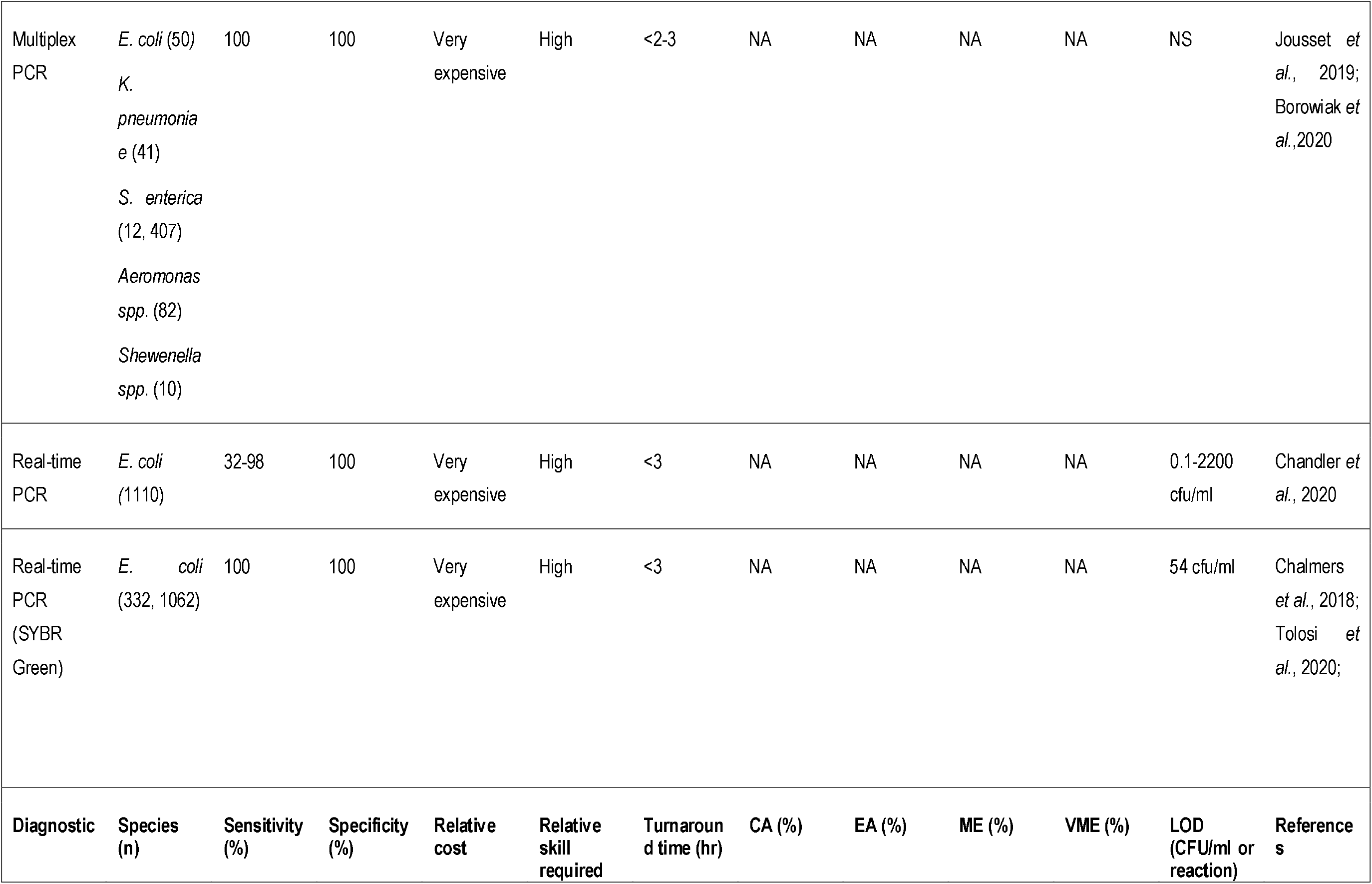
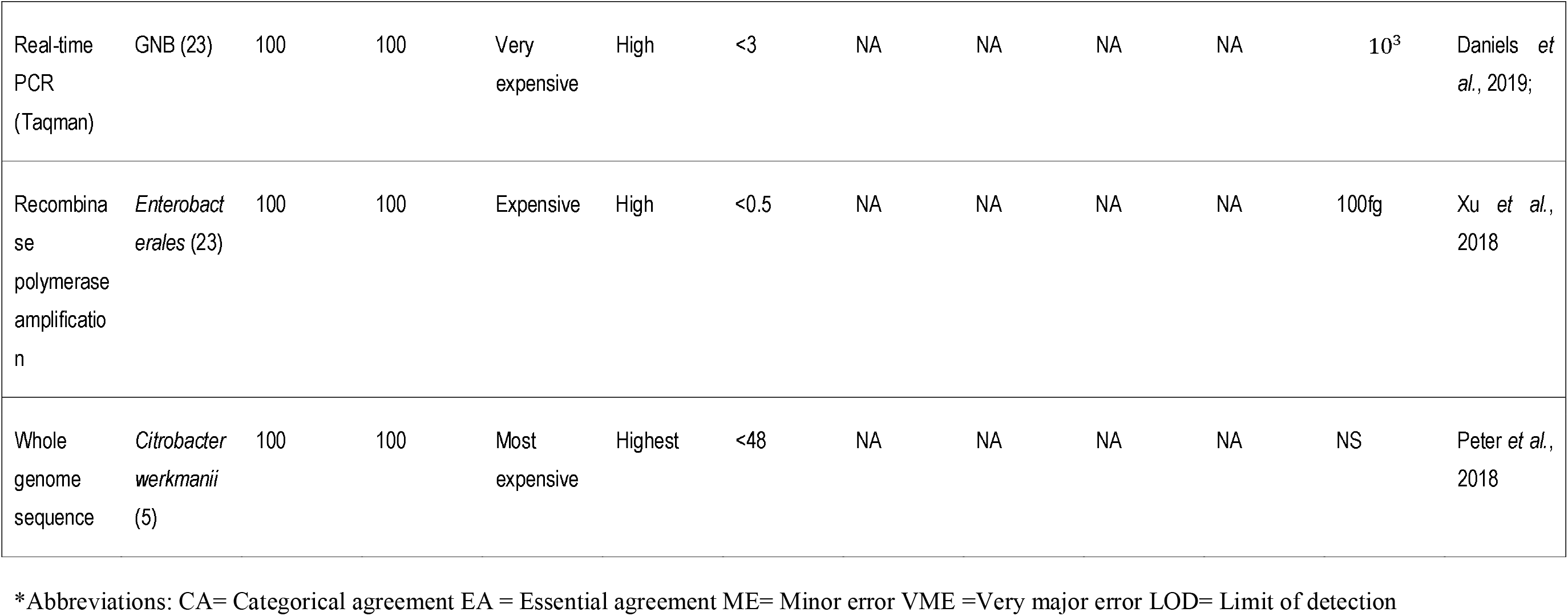
Comparative diagnostic efficiencies of colistin resistance diagnostics.

**Figure 1.**
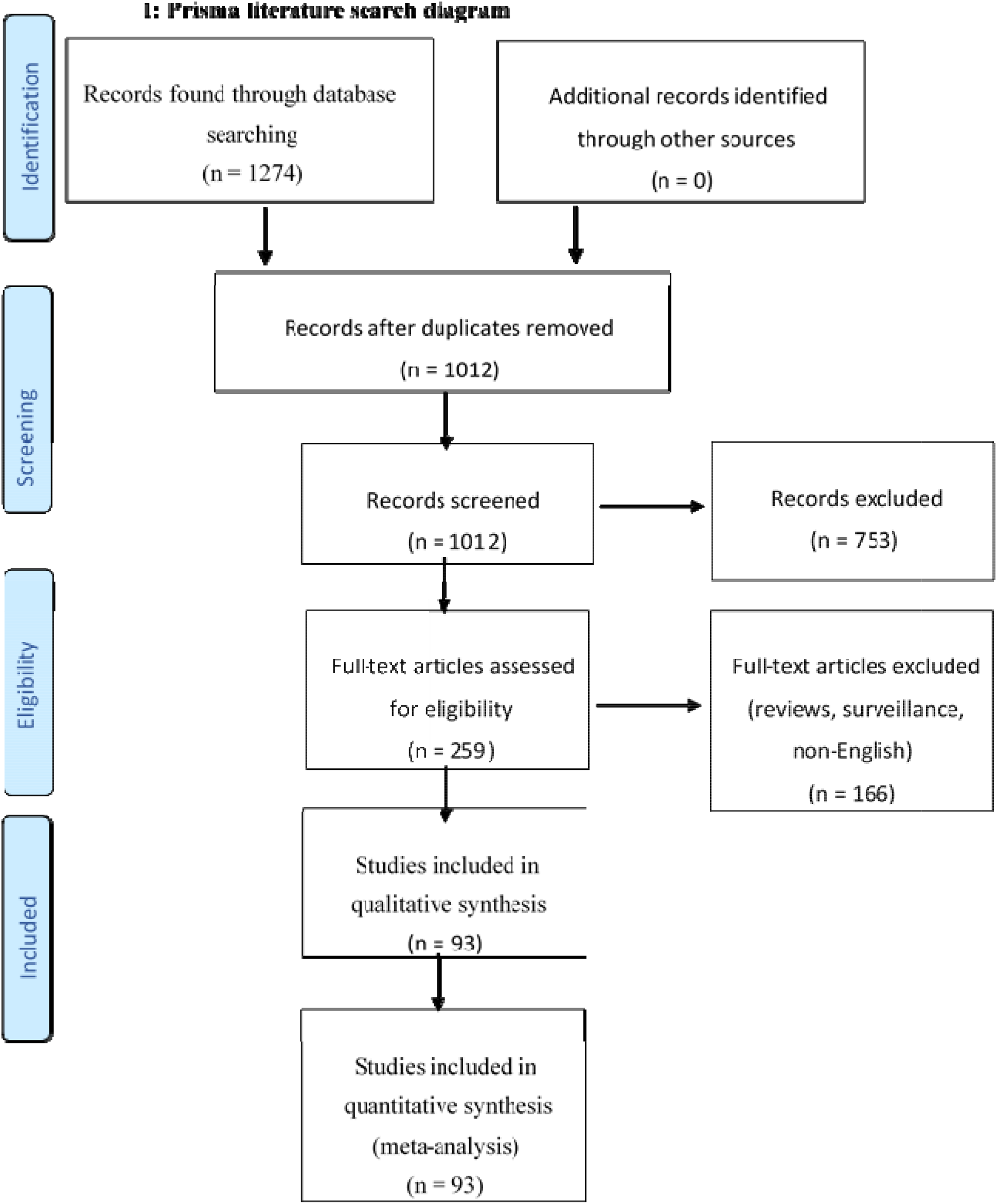
Literature search strategy and sorting techniques used to obtain articles for inclusion in the systematic review.

## 2. Phenotypic tests

### Broth Microdilution (BMD)

The Clinical and Laboratory Standard Institute (CLSI) and European Committee on Antimicrobial Susceptibility Testing (EUCAST) have jointly recommended that the MIC (minimum inhibitory concentration) testing of colistin be performed according to the ISO-20776 standard BMD method ^25, 26^. Methods such as agar dilution, disc diffusion and gradient diffusion have been ruled out as it was shown that the large molecular size of colistin makes it poorly diffusible through agar ^1, 26, 27^.

It has been recommended that BMD be used with plain polystyrene trays and colistin sulfate salt without the addition of any surfactants ^26, 28^. The CLSI had initially recommended the addition of polysorbate-80 (P-80) to alleviate the binding of polymyxin to negatively charged polystyrene surfaces (binding of colistin to polystyrene reduces the concentration of colistin)^1^. However, there were concerns that the surfactant would act in a synergistic manner with colistin, and is therefore not recommended at this time ^28^. Moreover, several studies have suggested that the loss of colistin concentration could be reduced by minimizing contact with unexposed pipette tips and by storing colistin solution in glass tubes ^28^.

The CLSI and EUCAST have established a colistin susceptible breakpoint of ≤ 2 mg/L and a resistant breakpoint of >2 mg/L for *Acinetobacter spp*. and *Pseudomonas aeruginosa* ^25^. However, only the EUCAST has the same breakpoints for *Enterobacterales* whilst the CLSI has an epidemiological cut-off value of 2 mg/L that defines *Escherichia coli, Klebsiella pneumoniae, Raoultella ornithiolytica, Enterobacter aerogenes* and *Enterobacter cloacae* as either wild type or non-wildtype ^1, 25^.

BMD performed by the reference ISO-20776 method is currently the only gold standard for determining colistin MIC and evaluating CA, EA, ME and VME; yet it is laborious and rarely performed in routine clinical microbiology laboratories ^1^. Instead, diffusion methods and automated AST systems are more commonly used ^1^. Chew *et al* (2017) evaluated the detection of *mcr*-1 positive *Enterobacterales* by BMD in comparison with commercial automated AST systems viz., Sensititre, Microscan and Vitek 2, and a gradient diffusion test *i.e*., E-test. This study found that none of the commercial testing methods meet the CLSI recommendation standard for commercial antimicrobial susceptibility testing systems: EA ≥90%, CA ≥90%, VME ≤1.5%, and ME ≤ 3.0%. Even so, the Sensititre and MicroScan were shown to detect 100% of the *mcr*-1 positive isolates (Refer to Automated AST systems below) ^25^. The BMD’s overall sensitivity could be improved by reducing the susceptible breakpoint to ≤1mg/L and using microtitre plates that were manufactured to reduce adsorption ^25, 28^.

The methodological challenges surrounding the standard BMD have led to interest in finding alternative means for detecting polymyxin resistance in Gram-negative bacteria^1, 28^. Broth macrodilution method has been shown to be an obvious alternative, as it employs the use of glass tubes instead of polystyrene ^28^. Notwithstanding, the broth macrodilution method is not a popular alternative because it requires the same preparation and TAT as the standard BMD. Hence, commercially available selective media and rapid colorimetric assays have become popular for screening (Table 1) ^28^.

### Diffusion and Agar dilution methods

Diffusion methods for colistin susceptibility testing are still commonly used despite being disapproved by the CLSI and EUCAST ^1, 27, 29, 30^. Two of the commercial gradient tests, E-test and MIC Test Strip, performed poorly for colistin-resistant isolates in one study ^30^. Disc diffusion is not an MIC determiner, although it perform similarly to gradient diffusion tests, generating high levels of VMEs (false-susceptible) whilst agar dilution has a tendency of generating higher MICs (which results in high MEs) than the reference method ^1, 25, 27, 28, 30-32^. These findings further support the conclusion that polymyxins are poorly diffused in agar and therefore corroborates the CLSI and EUCAST recommendations to abandon diffusion and agar dilution methods ^27, 30, 32^

### Manual commercial MIC-determiners: UMIC, MMS, and ComASP Colistin

UMIC (Biocentric, Bandol, France), MICRONAUT MIC-Strip (MMS) (MERLIN Diagnostika GmbH, Bornheim, Germany), ComASP Colistin (formerly SensiTest™, Liofilchem, Italy) are non-automated broth microdilution-based tests ^30^. UMIC and MMS both consist of a plastic device with 12 wells that allow for different colistin concentrations to be tested for a single isolate without the need for any additional equipment ^26, 30, 33^. Whilst the ComASP (SensiTest) Colistin, consists of a compact panel for four isolates with freeze-dried antibiotics in seven two-fold dilutions ^5, 34^. Matuschek *et al* (2018) evaluated all three tests, where ComASP had a poor EA for *Acinetobacter* spp., and UMIC was poor for *Acinetobacter* spp. and *P. aeruginosa*. The overall performance of ComASP was improved (as shown in Table 1) when certain species were removed and *K. pneumoniae* and *E. coli* were tested, suggesting that ComASP is not suitable for testing all species ^5^.

UMIC was generally found to lower the MIC of some colistin-resistant isolates, which may result in failure to detect colistin-resistant isolates with low MICs (≤8 mg/L) ^26, 33^. More so, UMIC failed to detect four *Stenotrophomonas maltophilia* isolates with MICs ranging from 8mg/L to 32mg/L in one study, although in another study all *S. maltophilia* isolates were detected accordingly ^26, 33^. The MMS had the best correlation to the BMD amongst the three tests; however, there aren’t sufficient studies evaluating this test (Table 1)^30^.

### Automated AST systems: BD Phoenix, MICRONAUT-S, MicroScan, Sensititre and Vitek 2

Automated antimicrobial susceptibility testing (AST) systems are of particular interest for colistin susceptibility testing due to their ease of use than the reference BMD ^26, 30^. Several studies have shown that some automated AST systems can achieve results that are relatively similar to those of the reference BMD ^25, 26, 30, 32, 35^. The Sensititre recorded a high rate of agreement with the reference BMD in several studies, particularly demonstrating the highest potential for detecting *mcr*-1-producing *Enterobacterales* together with MicroScan in one study ^25, 26, 30^. However, MicroScan has a tendency of overestimating MICs (which may result in false-resistant isolates) of *E. cloacae, Salmonella spp.* and non-fermenters (*A. baumannii, P. aeruginosa* and *S. maltophilia*) ^5, 26^. Vitek 2 had the highest rate of VMEs at 36% for *mcr*-1-producing isolates and failed to detect colistin resistance in eight *mcr*-negative *E. cloacae* complex isolates ^25^. However, another study found that Vitek 2 had VMEs of 2.04% for *K. pneumoniae* isolates that were *mcr*-negative ^36^. MICRONAUT-S (MERLIN Diagnostika GmbH, Bornheim, Germany) performed similarly to the Sensititre although it had two VMEs whereas the Sensititre had none ^30^. The Sensititre test is unique as it can be performed manually or semi-automated unlike MicroScan, MICRONAUT-S, Vitek 2 and BD Phoenix that require an automated inoculation delivery system ^26, 30^.

The detection of colistin resistance by BD Phoenix highly agreed with that of the reference BMD method ^32, 35^. Nevertheless, the BD Phoenix 100 system failed to detect colistin-resistant isolates that may have hetero-resistance; the BD Phoenix M50 had 5.7% MEs, all of which were within twofold dilutions, as compared to the reference BMD ^35, 37, 38^. Notably, the BD Phoenix M50 has not been evaluated on strains that exhibit heteroresistance; hence, its evaluation is limited compared to BD Phoenix 100 (Table 1) ^35^.

### Chelator-based and non-chelator-based tests

#### Rapid Polymyxin NP

The Rapid Polymyxin NP is a colorimetric test that is based on glucose metabolism to detect the growth of *Enterobacterales* at a given concentration of a polymyxin (colistin or polymyxin B) ^11^. Resistance to polymyxins is demonstrated by a colour change (orange to yellow) of a pH indicator, *i.e.* phenol red, as a result of acid formation associated with the metabolism of glucose ^11, 39, 40^. Rapid polymyxin NP test is commercially available (ELITechGroup, Puteaux, France) and can also be performed in-house with the preparation of two solutions ^11, 41^. The in-house rapid polymyxin NP test is prepared with stock solutions of polymyxins and a rapid polymyxin NP solution, which consists of cation-adjusted Mueller-Hinton broth powder, phenol red indicator and D(+)-glucose ^11^. This test has demonstrated an excellent detection of colistin resistance in several studies, including detecting colistin resistance directly from blood cultures with a sensitivity of 100% ^40, 42, 43^. However, in a study using ComASP as the reference, the Rapid Polymyxin NP test had a lower specificity than BD Phoenix, Vitek 2 and E-test with *K. pneumoniae* isolates (Table 1)^36^. In another study, Rapid Polymyxin NP test recorded a lower sensitivity and specificity (71.1% and 88.6%) than E-test, which had a sensitivity and specificity of 80.4% and 95.8% respectively ^44^. However, 10 isolates that were included in the calculations of the E-test performance were excluded from the Rapid Polymyxin NP test as they were considered non-evaluable due to no growth in the growth control ^44^.

Although the Rapid Polymyxin is limited to use on *Enterobacterales*, the test is easy to perform and the final results can be read in no more than four hours, with the majority of the results being positive in two hours ^31, 32, 42, 45^.

#### Andrade Screening Antimicrobial Test (ASAT)

Following the Rapid Polymyxin NP test, another colorimetric assay for detecting colistin resistance in *Enterobacterales* was developed ^13^. The Andrade Screening Antimicrobial Test (ASAT) was developed using an in-house broth consisting of 10g peptone, 5g sodium chloride, 3g beef extract and 10mL Andrade indicator made with 0.5g acid fuchsin and 16mL NaOH in 100mL water ^13^. The evaluation of the ASAT was performed on 300 *Enterobacterales* in tubes containing 175 µL of Andrade broth and colistin at a concentration of 3.8 mg/L ^13^. The test achieved a sensitivity and specificity of 90.7% and 100% respectively, where a positive result in the presence of colistin was shown by a change in colour of the Andrade indicator (light pink to magenta) ^13^.

The ASAT test was further evaluated against the BD Phoenix using 1096 *Enterobacterales* clinical isolates ^13^. However, this evaluation demonstrated discrepancies between the two methods where 10 *E. coli*, seven *K. pneumoniae* and one *E. cloacae* colistin-resistant isolates were accurately detected by ASAT and not by BD Phoenix ^13^. Most of the isolates that were not detected by BD Phoenix had colistin MIC values ranging between 4-8 µg/mL and carried *mcr*-1 genes ^13^. Although most of the colistin-resistant *K. pneumoniae* isolates had colistin MIC values >16 µg/mL and carried bla_KPC_genes, only three isolates with colistin MIC values ≤16 µg/mL showed discrepancies between the two methods ^13^.

#### Rapid Polymyxin Acinetobacter, Rapid Polymyxin Pseudomonas and Rapid Resapolymyxin NP

ElitechGroup introduced Rapid Polymyxin tests for *Acinetobacter spp*. and *Pseudomonas spp.* in October 2018 ^46, 47^. Both tests use the same principle as the Rapid Polymyxin NP test as they rely on the colorimetric detection of bacterial metabolism in the presence of a defined concentration of colistin ^47^. A positive result by Rapid Polymyxin *Acinetobacter* was read by a change in colour of a pH indicator, phenol red (red to orange or yellow), whereas the Rapid Polymyxin *Pseudomonas* uses bromocresol purple pH indicator (green-yellow to violet) ^47, 48^. Sadek *et al* (2020) evaluated the in-house version of the Rapid Polymyxin *Pseudomonas* test, which agreed with Lescat *et al* (2019), recording 100% sensitivity and a lower specificity of 95% ^47, 48^. However, the Rapid Polymyxin *Acinetobacter* had discrepancies, recording no errors in one study and eight errors (three VMEs and five MEs) in another ^46, 47^.

The Rapid Resapolymyxin NP test was also developed to detect colistin resistance in all colistin-resistant Gram-negative bacteria including *Enterobacterales, A. baumannii* and *P. aeruginosa* ^49^. The test was carried out by inoculating 20 µL standardized bacterial suspension (3.5 McFarland) in Mueller-Hinton broth containing a final concentration of 3.75 mg/L of colistin sulfate ^49^. A 10% concentration of resazurin PrestoBlue was added after three hours of incubating the medium and the results were read over a period of one hour after the addition of resazurin PrestoBlue ^49^. The detection of colistin resistance is based on the reduction of blue resazurin to pink resorufin by metabolically active cells in the presence of a defined concentration of colistin ^49^.

The evaluation of Rapid ResaPolymyxin NP test showed reliable detection of colistin resistance in non-fermenters and 100 % accuracy in *Enterobacterales* ^49-51^. Although the slow growing nature of *Pseudomonas* spp. resulted in an hour delay before a change in colour could be observed, *Enterobacterales* and *Acinetobacter* spp. were detected in less than four hours ^38, 50^. The Rapid ResaPolymyxin NP test makes up for the limitations of Rapid Polymyxin NP test in testing polymyxin resistance in Gram-negative bacilli (including non-fermenters) regardless of the mechanism of resistance ^49, 50^. Therefore, this test is more suitable for general categorization of colistin-resistant and colistin-susceptible isolates than the Rapid Polymyxin NP test ^50^.

#### EDTA/DPA-based colistin resistance tests

The MCR catalytic domain, phosphoethanolamine (PEtN) transferase, is a zinc metalloprotein, where zinc deficiency reduces colistin MICs in MCR-producing *E. coli* ^52-54^. The combined disk test, colistin MIC reduction test, modified rapid polymyxin NP test and alteration zeta potential, are four tests that are based on the inhibition of MCR activity by EDTA and have been strategically developed to detect *mcr*-genes ^52^.

The combined disk test (CDT) uses 10µL of 100mM EDTA solution, which is impregnated into one of two 10µg colistin disks ^52^. The two colistin disks are placed on Mueller Hinton agar plates swabbed with 0.5 McFarland bacterial suspension and incubated for 18 to 24h at 37[^52^. Results are read as positive if there is an increase of ≥3mm in inhibition zone around the colistin disk containing EDTA as compared to the colistin disk without EDTA ^52^. This method has recorded a sensitivity and specificity of 96.7% and 89.6% respectively by the developer ^52^. However, a latter study recorded less promising results, with a sensitivity and specificity of 12% and 65.2% respectively, Hence, further evaluation studies must be conducted on this test as the current results seem unreliable ^54^.

A pre-diffusion method of the CDT was evaluated, where two colistin disks were placed and allowed to diffuse for 2 hours on MH agar ^53^. The disks were removed and the plates were left at room temperature for 18-24h after which two disks containing 1µmol of EDTA were strategically placed (one exactly where the colistin disk had been placed and the other, at least 30 mm away) ^53^. Diameters of inhibition zones were measured after 18h of incubation and colistin-resistant MCR-positive isolates demonstrated a ≥5mm increase in inhibition zone around the disks ^53^. The pre-diffusion method, using a cut-off value of ≥5mm, improved the CDT to 100% accuracy for *mcr* detection ^53^; it has however not been extensively evaluated to ensure reproducibility and seems more complicated, laborious and time-consuming than the CDT.

Colistin MIC reduction (CMR) test was performed by broth microdilution using MH broth without cation supplementation but with 80µg/mL EDTA solution instead ^52^. It was considered that cation supplementation with calcium and magnesium would impair the inhibitory activity of EDTA; moreover, calcium could favour the activity of putative PEtN transferases in *E. coli* ^52^. Even so, this method did not efficiently detect *mcr*-producers among *Enterobacterales* isolates although different concentrations of EDTA were used ^52, 55^.

Innovatively, EDTA has been added to the Rapid Polymyxin NP test in what is termed the modified rapid polymyxin NP (MPNP) test to enable it to identify MCR producers. MPNP is the Rapid Polymyxin NP test with the addition of two wells filled with colistin-free solution and colistin-containing solution, both with 80µg/mL EDTA ^52^. Results were read as positive for the production of MCR-1 PEtN transferase if there was no change in colour of red phenol in the presence of colistin and EDTA ^52^. The presence of EDTA in the MPNP test successfully detected MCR-1 positive colistin-resistant *E. coli* isolates as demonstrated by a sensitivity and specificity of 96.7% and 100% respectively ^52^.

Finally, addition of EDTA results in an alteration of zeta potential of membrane charge, which is measured to determine MCR expression ^52^. Particle size and zeta potential of colistin-susceptible and colistin-resistant bacterial cells grown in MH broth with or without 80µg/ml EDTA was measured using a ZETAPALS zeta potential analyser ^52^. Colistin-susceptible and colistin-resistant MCR-1 *Enterobacterales* demonstrated zeta potential values between -21.54 and -44.21 mV whilst colistin-resistant MCR-1 positive had ≤-20 mV (-4.20 to -19.34 mV) ^52^. In the presence of EDTA, an alteration of zeta potential ranging from -21.13 to -40.81 was observed in colistin-resistant MCR-1 positive *E. coli* isolates ^52^. A zeta potential ratio (*R*_*zp*_ = *ZP*_*+EDTA*_ /*ZP*_−*EDTA*_) was calculated for all isolates and a cutoff value of *R*_*zp*_ ≥2.5 as a criterion for the presumed detection of MCR-1-positive *E. coli* isolates was established ^52^. Alteration of zeta potential yielded a sensitivity and specificity of 95.1% and 100% respectively. However, EDTA had no inhibitory effect on *mcr*-1-positive *K. pneumoniae* isolates ^52^.

#### Colistin broth-disk elution (CBDE)

Simner *et al* (2019) developed the colistin broth-disk elution method (CBDE), which was performed on a collection of *Enterobacterales, P. aeruginosa* and *A. baumannii* isolates: four tubes were assigned to each isolate ^56^. The four tubes contained 10mL of CA-MHB with colistin disks to yield final concentrations of 0,1,2 and 4 µg/mL, respectively ^56^. The tubes were incubated for 30 minutes at room temperature, allowing colistin to dissolve into the broth, after which 50µL aliquot of 0.5 McFarland standard bacterial suspensions were added to each tube ^56^. Colistin MIC values were visually read after 16-20h of incubation at 35[in ambient air ^56^. In this study, CBDE was compared to the reference BMD and Sensititre, where the CBDE had a CA and EA of 98% and 99%, respectively as compared to both BMD methods ^56^. Three *mcr*-1-producing *E. coli* isolates resulted in a VME rate of 8% due to one dilution difference by CBDE and BMD; however, no errors were observed when CBDE was compared to BMAD ^56^.

Three studies have evaluated the modified version of the CBDE ^57-59^. Bell *et al* (2019) was the first to describe a modified CBDE method. The method was performed as previously described, however 1mM EDTA was used by adding 20µL of 0.5M EDTA to each tube containing CA-MHB and 10µg colistin disks ^58,59^. Fenwick *et al* (2020) added a fifth tube to generate CBDE+EDTA with colistin concentrations of 0, 0.4, 1, 2 and 4 µg/mL, respectively. The CBDE+EDTA method has shown overall sensitivity and specificity of 100% and 94.3-95.8% respectively, for screening the presence of MCR in *Enterobacterales* and *P. aeruginosa* ^58, 59^.

Dalmolin *et al* (2019) evaluated the CBDE method using final volumes of 1mL and 200µL in colistin broth microelution (CBM) and the microelution test (MPT), respectively. The two methods were evaluated on 68 *Enterobacterales*, nine *A. baumannii* and eight *P. aeruginosa* isolates from human and animal samples ^57^. Both CBDE methods were performed as previously described, however the CBDE mixture was fractioned in 1mL tubes for the CMB test and 200µL in microtiter plates for MPT test ^57^. Additionally, this study evaluated the colistin susceptibility test tube (CSTT), which was performed using one tube with 5mL CA-MHB and 10 µg colistin disk to yield a final concentration of 2 µg/mL ^57^. All three methods presented unsatisfactory MEs and VMEs; particularly, they performed poorly for non-fermenters ^57^.

Colistin broth-disk elution methods are performed using reagents that are readily available at low cost ^56, 59^. However, CBDE with EDTA could be more suitable for screening *mcr*-positive isolates as CBDE alone tends to underestimate MICs of *mcr*-positive isolates ^56, 58, 59^.

#### Colistin-MAC test

*The colistin-MAC test was designed to detect* mcr*-genes on the basis of colistin MIC reduction by a fixed concentration (900µg/mL) of dipicolinic acid (DPA) ^60^. The test was carried out in 96-well microtitre plates using CA-MHB with DPA stock solution prepared in dimethyl sulfoxide ^60^. Coppi* et al *(2018) established a cutoff value of* ≥*8-fold colistin MIC reduction in the presence of DPA for the presumptive identification of* mcr*-positive isolates. However, a latter study used a different cutoff value, where* ≥*3 twofold MIC reduction in the presence of DPA indicated a positive result ^55^. The Colistin MAC test was found to perform well for* E. coli *isolates and ineffective in detecting* mcr*-genes of* K. pneumoniae *and* Salmonella *spp. ^5, 55, 60^. The lack of inhibitory effect of DPA in* K. pneumoniae *isolates can be attributed to a decrease in DPA permeability or the existence of other mechanisms of colistin resistance in these strains ^60^. Moreover, the addition of DPA resulted in reductions and increments in MICs of some isolates, although these adjustments did not affect the accurate sensitivity classification of the isolates ^5^. Colispot*

Colispot is a test developed by Jouy *et al* (2017), it consists of applying a single drop of 8 mg/L colistin solution on MH agar to detect colistin resistance. The test was initially carried out by applying 10µL drop of colistin (twofold concentrations ranging from 0.25 to 256 mg/L) on MH agar inoculated with 10^5^ *E. coli* suspensions ^61^. Each drop was strategically placed so that their centres were at least 2 cm away from each other to allow for an inhibition zone of >5 mm ^61^. The colispot test was evaluated on 106 *E. coli* isolated from veterinary faecal samples and 35 *mcr*-1 positive *E. coli* isolates from bovine samples ^61^. Susceptible isolates had a clear inhibition zone around colistin drops with concentrations ranging from 0.25 to 4mg/L although the size of inhibition zone was dependent on the colistin concentration tested ^61^. A clear inhibition zone of 8 to 10 mm was observed with all the susceptible isolates when a single concentration of colistin solution with CLSI/EUCAST bacterial inoculum size and incubation temperature were used ^61^. This test can be routinely used when performing diffusion methods as it can improve the detection of acquired resistance in *E. coli* ^61^.

### Lateral Flow-Immunoassay

Monoclonal antibodies (MA) were used to develop lateral flow assays to detect MCR-1-producing bacterial isolates ^62^. In this study, 177 and 121 *Enterobacterales* isolates from human and animal samples were obtained, respectively ^62^. Bacterial colonies were isolated from agar plates, suspended in extraction buffer and dispensed on the MA-containing cassette where they were allowed to migrate for 15 minutes ^62^. All MCR-1-producing isolates were detected accordingly as shown by a pink band on the test line and control line of the assays. Furthermore, this test was able to detect MCR-2-producing isolates ^62^. The same assay is currently marketed as the NG-Test MCR-1 by NG Biotech in France ^58^. Initially, the evaluation of the NG-Test MCR-1 resulted in eight false-positive results that were ultimately resolved to negatives apart from one isolate that was found to be an MCR-2 producer ^58^. Whilst the detection of the MCR-2 product by the NG Test MCR-1 further confirms the results of the developers, cross-reactivity with MCR-2 limits the accuracy of the assay for the MCR-1 producers. ^58^. Nonetheless, the lateral flow immunoassays were found to be highly sensitive, easy to use and cost-effective for detecting MCR-1/-2 ^58, 62^.

### Agar-based screening medium

#### Superpolymyxin

Superpolymyxin is a selective medium for polymyxin-resistant Gram-negative bacteria that is based on eosin methylene blue agar ^63^. The medium was developed with the optimal colistin concentration of 3.5 µg/mL, 10 µg/mL of daptomycin (to inhibit potential growth of Gram-positive strains) and 5 µg/mL of amphotericin B as an antifungal. Nordmann *et al* (2016) designed Superpolymyxin for screening intrinsic and acquired polymyxin-resistant Gram-negative bacteria as previous screening media containing deoxycholic acids and a high concentration of colistin inhibited the growth of strains with acquired resistance and low MIC values (Table 1).

The use of eosin Y and methylene blue dyes helped distinguish lactose-fermenters (dark brown to purple) from non-fermenters (colourless) ^11^. This medium distinguishes lactose-fermenting *E. coli* (metallic green sheen) from other *Enterobacterales*, including non-fermenting *E. coli* (dark brown to purple) ^64, 65^. However, studies evaluating other selective media against Superpolymyxin have shown a weaker detection of non-fermenters by Superpolymyxin ^63, 66^.

This medium was able to detect colistin-resistant Gram-negative bacteria directly from bacterial culture and clinical samples (*i.e.,* rectal swabs and stool samples) with high sensitivity and specificity ^44, 67, 68^. However, direct inoculation from clinical swabs may result in the growth of colistin-susceptible isolates on the medium, therefore resulting in a poor specificity (as low as 80.45%) ^44, 68^. The poor specificity of Superpolymyxin for clinical samples was suspected to be due to sample storage conditions and bacterial inoculum effect (≥10^6^ CFU/mL) ^44, 67^.

Two studies have recorded a low sensitivity (≤77.3%) for *Enterobacter* spp., which may be due to hetero-resistant phenotypes (*i.e.*, may have a small population of bacterial cells with colistin resistance) ^67, 69^. In both studies, the Superpolymyxin plate was inoculated with 10 µL of a 0.5 McFarland bacterial suspension. Therefore, a higher inoculum for *Enterobacter* spp. was suggested ^67, 69^.

#### CHROMagar™ COL-APSE

CHROMagar™ COL-*APSE* by CHROMagar (Paris, France) is the first selective medium designed to detect and differentiate all *Acinetobacter spp*., *Enterobacterales, Pseudomonas spp*. and *Stenotrophomonas spp*. ^63^. The agar plates were prepared in-house using a dehydrated CHROMagar base medium and supplements (S1 and X192) containing colistin sulfate and oxazolidinones to enhance the growth of colistin-resistant Gram-negative bacteria and inhibit that of Gram-positive bacteria ^63^. Swarming by *Proteus spp.* was inhibited by adding p-nitrophenyl glycerol to the medium preparation, which did not disrupt the medium’s performance. This makes CHROMagar™ COL-*APSE* suitable for screening mixed specimens^63^.

The accuracy in detecting and differentiating colistin-resistant Gram-negative species was evaluated by Osei Sekyere *et al.* (2020), where the morphological appearance of the detected strains was as described by the manufacturer. Moreover, three studies that have evaluated CHROMagar™ COL-*APSE* agreed that the medium had a high sensitivity in detecting isolates harbouring *mcr* genes ^5, 63, 70^. However, there was significant difference in the sensitivity and specificity recorded by Abdul Momin *et al* (2017) and Osei Sekyere *et al* (2020) (Table 1). The poor performance in the recent study could be due to the use of cultured bacteria instead of using serial dilutions in broth (Table 1)^5, 63^.

#### ChromID® Colistin R

ChromID® Colistin R is a chromogenic selective medium that is primarily used for isolating colistin-resistant *Enterobacterales* from clinical stools and rectal swab samples ^71^. Similar to CHROMagar™ COL-*APSE*, the medium can differentiate between bacterial species based on morphological appearance of bacterial colonies *i.e., E. coli* (pink to burgundy), *Klebsiella spp., Enterobacter spp., Serratia* (blue to green), *Salmonella spp*. (white or colourless) and *Proteeae* tribe (beige-brown) (Table 1) ^64, 71^.

The assessment of ChromID® Colistin R and Superpolymyxin using stool and rectal swab samples resulted in an overall better performance by ChromID® Colistin R ^64^. The lower limit of detection (LOD) of this medium being at least one log lower in 69.2% of the isolates detected on both media, whereas Superpolymyxin only had a better LOD for 7.7% isolates ^64^. Nonetheless, Superpolymyxin could be directly inoculated with stool or rectal swab samples without a 4-5 hours enrichment step required by ChromID® Colistin R; and the final sensitivity (84.9 to 86.8%) recorded for this medium was achieved after extending the TAT from 24 hours to 48 hours (which also allowed for the detection of an *mcr*-1 producing *E. coli* isolate) ^64^.

A study by Thiry *et al.* (2019) evaluated this medium against CHROMagar™ COL-*APSE* on 158 colistin-resistant bovine *E. coli* isolates. Half (48/96) of the isolates considered to be intermediate to the disk diffusion test had MIC > 2 and were able to grow on both media, with two more isolates growing on ChromID® Colistin R alone ^70^. Although both media could support the growth of (21/22) *mcr*-1-positive and (13/14) *mcr*-2-positive isolates, CHROMagar™ COL-*APSE* has an advantage over ChromID® Colistin R as it is not limited to (isolating and differentiating) *Enterobacterales* (Table 1) ^63, 70^

#### LBJMR medium

Lucie-Bardet-Jean-Marc-Rolain (LBJMR) medium, a polyvalent medium based on Purple agar, has been designed for the isolation of colistin-resistant Gram-negative bacteria as well as vancomycin-resistant Gram-positive bacteria ^66^. The medium was developed by adding glucose (7.5 g/L), colistin sulfate (4 μg/mL) and vancomycin (50 μg/mL) to 31g/L Purple agar base ^66^. One hundred and forty three bacterial isolates, including colistin-resistant *Enterobacterales* and non-fermentative Gram-negative bacilli were used to evaluate this medium, where the specificity and sensitivity were 100% ^66^. The medium was further evaluated on 56 *mcr*-1 positive and 10 *mcr*-1-negative chicken and human stool samples as well as two clinical rectal swabs ^66^. The study found that the LBJMR could detect *mcr*-1 positive isolates with high sensitivity, particularly showing a higher sensitivity for colistin-resistant non-fermenters than Superpolymyxin ^66^. Furthermore, the LBJMR medium does not contain daptomycin and amphotericin B, which are used in some of the agar-based media *i.e.* Superpolymyxin, to inhibit the growth of Gram-positive bacteria, including vancomycin-resistant *Enterococcus* ^66^

#### Luria-Bertani (LB) medium

A selective medium for detecting colistin-resistant *Enterobacterales* (including those with *mcr*-1 genes) in spiked stools was evaluated ^72^. The medium was developed by adding 4mg/mL colistin, 10mg/mL vancomycin, and 5mg/mL amphotericin B to agar medium made with 25g of Luria-Bertani (LB) powder ^72^. Nine *Enterobacterales* isolates were collected, seven of which carried *mcr*-1 genes ^72^. Each of the nine isolates were spiked into donated faecal samples and serially diluted to final concentrations of 10^2^ or 10^3^ CFU/mL; 0.5 mL of the stool mixture were spiked in 4.5 mL of *Enterobacterales* enrichment (EE) broth and incubated for 24h at approximately 35□ ^72^. Afterwards, 10 µL of the spiked EE broth was inoculated onto the LB medium and incubated at 35□±2□ for 48 hours in ambient air ^72^. The selective LB medium demonstrated a sensitivity of 100% (Table 1)^72^.

## 3. Molecular tests

### Amplidiag Carba-R+ MCR assay

The Amplidiag Carba-R+ MCR assay is a multiplex nucleic acid-based test developed for detecting carbapenemase and *mcr*-1/-2 genes from rectal swabs and bacterial culture ^73^. The assay was performed retrospectively and prospectively on 215 Gram-negative bacilli and 51 *Enterobacterales* isolates, respectively ^73^. The Amplidiag Carba-R+MCR assay did not detect one GES carbapenemase-producing *P. aeruginosa.* However, all *mcr*-1 and *mcr*-2-producing isolates were accurately detected (Table 1)^73^.

Additionally, this assay was performed on DNA extracted from 100 rectal swabs, including 40 carbapenemase-positive samples: the sensitivity was 92.5% ^73^. Two NDM and one OXA-48 producers were not detected due to a low concentration of bacteria; therefore, the samples were subjected to an overnight enrichment in brain heart infusion with 0.5 µg/mL ertapenem ^73^. The enrichment step allowed for the detection of two of the three samples that were not previously detected. One of the samples was an OXA-48 producing *E. cloacae* with an AcOXA (*Acinetobacter* oxacillinases with carbapenemase activity) gene ^73^. The overall performance of this assay was acceptable, demonstrating sensitivity and specificity ranging from 92-100% and 86-100% respectively ^73^. Moreover, the assay can be performed on cultured bacteria as well as DNA extracted from rectal swabs in no more than three hours (Table 1)^73^.

### AusDiagnostic MT CRE EU assay

AusDiagnostics MT CRE EU assay is a two-step nested multiplex-tandem PCR (MT-PCR) assay by AusDiagnostics (Chesham, UK) ^74^. One study evaluated the AusDiagnostics MT CRE EU assay for detecting carbapenemase, *mcr*-1, and *mcr*-2 genes ^74^. A collection of *Enterobacterales, Pseudomonas spp.* and *Acinetobacter spp*., including carbapenemase or *mcr*-1/-2 producers, were used to evaluate the performance of this assay ^74^. The assay was performed by suspending two to three bacterial colonies grown overnight on Columbia blood or cystine lactose electrolyte agar in tubes with a sample buffer ^74^. The tubes were loaded onto the AusDiagnostics MT processor platform for template extraction and the first round of PCR ^74^. Lastly, the nested RT-PCR was performed by loading a 384-PCR plate containing the reaction mix onto the AusDiagnostic MT analyser ^74^. The results were automatically read using the AusDiagnostics MT assay software. The assay failed to detect four out of the 22 *mcr* genes; however, the *mcr* genes were also not detected by the reference PCR ^74^.

Retrospective and prospective evaluation of the assay resulted in eight and 18 false-positive results, respectively ^74^. An overall sensitivity and specificity of 95.5% and 99.8%, respectively, were obtained, which improved to 100% following repeats of the assay ^74^. The AusDagnostic MT CRE EU assay detected *mcr*-1/-2 genes as well as carbapenemase genes with minimal hands-on time (Table 1)^74^.

### Loop-mediated isothermal amplification (LAMP)

Loop-mediated isothermal amplification (LAMP) is a nucleic acid amplification method that allows autocycling strand displacement DNA synthesis at constant temperature using Bst DNA polymerase ^75^. The use of LAMP for detecting *mcr*-1 gene was first described by two studies ^76, 77^. Zou *et al* (2017) established a LAMP assay for to detect *mcr*-1 gene from cultured bacteria and spiked human stools. In this study, the LAMP assay was performed in 25 µL reaction mixtures that contained 20mM Tris-HCl, 10mM KCl, 10mM (NH4)_2_ SO_4_, 8mM MgSO_4_, 0.8 M betaine, 0.1% Tween-20, 1.4 mM of each DNTP and 8U Bst DNA polymerase ^77^. Each reaction mixture had specified amounts of forward and backward inner primers, outer forward and backward primers, loop primers as well as the appropriate amount of DNA template ^77^. The mixture was covered with 25µL wax and incubated in a dry bath incubator for 60 minutes at constant temperature, the amplification products were read visually and by turbidimetry ^77^. Visual detection was by colour change of a fluorescent metal indicator, a positive reaction was demonstrated by the formation of a magnesium pyrophosphate precipitate, which changed the reaction mixture from orange to green during amplification ^77^.

Imirzalioglu *et al* (2017) evaluated the eazyplex SuperBug *mcr*-1 kit (Amplex Biosystems GmbH, Giessen, Germany) for the rapid detection of *mcr*-1 gene. Colistin-resistant isolates were grown on LB media containing colistin sulfate to prevent the loss of *mcr*-1 encoding plasmid, whilst colistin-susceptible isolates were grown in the absence of colistin ^76^. The *mcr*-1-detecting LAMP assays were found to detect *mcr*-1 genes accurately, and the in-house LAMP assay was stated to be more sensitive than conventional PCR assays (Table 1) ^76, 77^.

However, *mcr*-1 detecting LAMP assays cannot detect other potential target genes ^75-77^. Therefore, Zhong *et al* (2019) developed a restriction endonuclease-based multi-LAMP for the detection of multiple *mcr*-genes. Two separate LAMP systems were established, a double-LAMP (*mcr*-2 and *mcr*-5) and triple-LAMP system (*mcr*-1, *mcr*-3 and *mcr*-4) which were performed in 25 µL reaction mixtures ^75^. The 25 µL reaction mixture consisted of 12.5 µL LAMP-reaction mix, 1 µL Bst 2.0 polymerase, 1.25 µL primer mix, 8.25 µL nuclease-free water, and 2 µL DNA lysate ^75^. Amplification products were detected visually by change in colour of SYBR Green I, which changed from yellow to orange for a positive reaction ^75^. Amplification products were also stained with GoldView TM and analysed by electrophoresis on 2% agarose gel ^77^. Multiplex detection of *mcr*-1 to *mcr*-5 genes was established through restriction digestion of the LAMP products based on band numbers and fragment lengths using Hind restriction enzyme ^75^.

LAMP assays have advantages over conventional PCR in that LAMP is more sensitive, has a shorter processing time of <60 minutes, is relatively easier to run, and multiplex detections can be conducted in the same detection system (Table 1) ^75-77^.

### Matrix-assisted laser desorption ionization-time of flight mass spectrometer (MALDI-TOF MS)

MALDI-TOF MS is a technique that is based on the production of mass spectra from whole cells and their contrast to a reference spectrum ^78^. This method is widely used for species identification of pathogens in clinical microbiology laboratories. Giordano and Barnini were the first to evaluate the possibility of detecting colistin resistance using MADI-TOF MS ^78^. In this study, 139 *K. pneumoniae* isolates from clinical samples were collected, from which protein was extracted for identification using MALDI-TOF MS (Bruker Daltonik GmbH, Bremen, Germany)^78^. The MICs of the isolates were determined by Sensititre using a range of antibiotics including colistin ^78^. First, a training set for mass peak analysis was established using 50/139 of the *K. pneumoniae* isolates from which 400 spectra were obtained and used for database entry (Main Spectrum Profile) as well as to classify algorithm models ^78^. Finally, the remaining 89/139 isolates were used to conduct the test; 712 spectra were collected from this set ^78^. However, from the 712 spectra, 158 were excluded as they constituted flat-line spectra or outliers, demonstrating identification score below 2.3 ^78^. Based on the mass signals and intensities of the bacterial protein samples, two-dimensional peak distribution classified the training set spectra into two main groups viz., colR-KPn (colistin-resistant *K. pneumoniae*) and colS-KPn (colistin-susceptible *K. pneumoniae*) isolates ^78^.

The newly created database entry consisted of using *MALDI Biotyper RTC* and *MALDI Biotyper* v3.0 to identify *K. pneumoniae* isolates and for the automatic detection of colistin resistance, respectively ^78^. The automatic classification of the test set resulted in the correct classification of 71% colR-KPn and 40% colS-KPn ^78^. Furthermore, different algorithm models were tested using ClinProTools v3.0 (Bruker Daltonics). The three algorithms tested included the Genetic Algorithm (GA), Supervised Neural Network (SNN) and Quick Classifier (QC)^78^. However, the tested algorithms either had good recognition capability and cross validation but poor classification of colistin resistance or poor recognition capability and acceptable classification of colistin resistance ^78^. The GA seemed more promising as it was better suited for biological samples ^78^.Therefore, a reliable classification model was created by combining the most relevant peaks detected from the GA algorithm^78^. The resulting peak combination of 4507.28/5142.84 Da from GA demonstrated a sensitivity and specificity of 78% and 89%, respectively ^78^.

Three studies have evaluated MALDIxin, a MALDI-TOF-based assay, ^7, 22, 79^. The MALDIxin test was developed to detect pETN modification in lipid A directly from bacterial colonies in less than 15 min ^22, 79^. Dortet *et al* (2018) evaluated MALDIxin on *A. baumannii* isolates, where the mass spectrum in colistin-susceptible isolates was characterised by two sets of peaks at the centre of *m/z* 1728.1 and *m/z* 1910.3. The peaks were assigned to bis-phosphorylated hexa-acyl and bis-phosphorylated hepta-acyl lipid A that had 12 to 14 carbons making up the acyl chain, respectively ^79^. The mass spectrum in colistin-resistant isolates was observed by two sets of peaks at the centre of *m/z* 1935.3 and *m/z* 2033.3, showing m/z+25 and m/z+123 shifts of mass unit of the bis-phosphorylated hepta-acyl lipid A at *m/z* 1910.3 ^79^. The peaks observed at *m/z* 2033.3 and *m/z* 1935.3 were assigned to pETN-modified-bis-phosphorylated hepta-acyl and pETN-modified-mono-phosphorylated hepta-acyl lipid A, respectively with an acyl chain of 12 carbons in length ^79^. The peaks (*m/z* 2033.3 and *m/z* 1935.3) associated with pETN-modified lipid A were observed in all colistin-resistant isolates and were not observed in any of the colistin-susceptible isolates ^79^.

Furniss *et al* (2019) and Dortet *et al* (2020) described the optimization of the MALDIxin test for detecting colistin resistance in clinical *E. coli* and *K. pneumoniae* isolates, respectively. Furniss *et al* (2019) optimized the MALDIxin test by adopting the low-resolution linear mode used by the MALDI Biotyper Sirius system. The optimization was achieved by adding a mild-acid hydrolysis step, which is required for analysis of clinical isolates in negative ion mode ^7^. The mild-acid hydrolysis step was performed by resuspending a single bacterial colony grown on MH agar for 18-24h in 200µL distilled water ^7, 22^, after which 50-100 µL of 2% acetic acid was added to double-distilled water containing bacterial suspension and heated for 5-15 min at 98-100□^7, 22^. For MALDI-TOF analysis, Furniss *et al* (2019) used a MALDI Biotyper Sirius system, whereas Dortet *et al* (2020) used a 4800 Proteonic Analyzer. The optimization of the MALDIxin allowed for the identification of L-Ara4N-and pETN-modified lipid A in *E. coli* and *K. pneumoniae* isolates. Moreover, the optimized methods were able to distinguish between chromosome-encoded and MCR-mediated colistin resistance ^7, 22^.

### Microarray

A commercial CT103XL microarray system that allows for the simultaneous detection of *mcr*-1*/-*2 and β-lactamase genes was evaluated ^80^. The study was conducted on 106 *Enterobacterales* isolates including *mcr*-1 and *mcr*-2 positive strains, as well as carbapenemase and extended-spectrum β-lactamase (ESBL)-producing strains ^80^. The CT103XL microarray, which uses a multiplex ligation detection reaction, was performed following bacterial DNA extraction from bacterial cultures ^80^. The commercial CT103XL microarray was confirmed to simultaneously detect *mcr*-1/-2 and β-lactamase genes with accuracy, although it failed to detect *mcr*-3, which shares 45% and 47% identity to *mcr*-1 and *mcr*-2, respectively ^80^.

### Multiple Cross Displacement Amplification coupled with Gold Nanoparticles-Based lateral Flow Biosensor

A multiple cross displacement amplification (MCDA) method, coupled with gold nanoparticles-based lateral flow biosensor (LFB) assay for detecting *mcr*-1 gene was developed ^81^. The MCDA reaction was performed on extracted DNA from 59 bacterial isolates, where each 25 µL reaction consisted of 12.5 µL reaction buffer, 1µL Bst DNA polymerase 2.0, 1µL colorimetric indicator, 1.6 µM of each cross primers, 0.4 µM of each displacement primers, 0.4 µM amplification primers and 1 µL DNA template ^81^. The MCDA reaction systems were then subjected to isothermal temperature (63□) for 40min, after which the amplification products were analysed using 2% agarose gel electrophoresis, colorimetric indicator and LFB ^81^. For *mcr*-1 detection by LFB, 0.2 µL of the amplicons was added to the well of the sample pad, followed by the addition of three drops of running buffer (1% Tween 20 and 0.01 mol/L phosphate-buffered saline) ^81^. The results were visually read after 1-2 min; a positive result was demonstrated by two red bands, one at the test-line and the other at the control line ^81^. The results were positive for all *mcr*-1 positive isolates and negative for all non-*mcr*-1 isolates ^81^.

The MCDA-LFB assay was further applied to stool samples spiked with 100 µL dilutions of bacterial strains ^81^. The resulting detection limit was 600fg of *mcr*-1 plasmid DNA per microliter in bacterial culture and 4.5 10^3^ CFU/mL in the spiked faecal samples ^81^. The MCDA-LFB has demonstrated the same sensitivity as the *mcr*-1 LAMP, which is more sensitive than the conventional PCR. Further, the MCDA-LFB has demonstrated a shorter reaction time ^77, 81^.

### Recombinase polymerase amplification (RPA)

The rapid detection of *mcr*-1, using a RPA, has been described ^82^. RPA is a novel isothermal amplification method, which can be performed in no more than 30 minutes at body temperature without the need for thermal cycling instruments ^82^. This study used basic RPA (B-RPA) and RPA with lateral flow (LF-RPA) on 23 genomic DNA extracted from 20 *mcr*-1 positive and three *mcr*-1-negative *Enterobacterales* ^82^. The primers for the B-RPA assay were designed by Clone Manager 8 and validated on three *mcr*-1 positive and one *mcr*-1 negative DNA samples ^82^. The B-RPA was based on the TwistAmp Basic kit reaction system, which was incubated at room temperature for 30 min, after which the amplicons were extracted by phenol/chloroform solution or purified using an amplicon purification kit ^82^. The LF-RPA reaction required primers and a probe, which were labelled with biotin and fluorescence ^82^. The LF-RPA was based on TwistAmp Nfo kit reaction system, which was incubated as described for the B-RPA ^82^. The amplification products for the LF-RPA were diluted at 1:50 with running buffer, after which a downstream operation was carried out ^82^.

The results for the B-RPA assay were read by agarose gel electrophoresis, whereas the results for the LF-RPA assay were visually read using Hybridetect 2T dipsticks ^82^. A positive *mcr*-1 detection by LF-RPA was demonstrated by two purple bands at the test line and the quality control line ^82^. Both assays detected the *mcr-*1 positive and *mcr*-1 negative DNA samples accordingly; therefore, both assays are equally suitable for detecting *mcr*-1 genes ^82^.

### Conventional & real-time PCR and Whole genome sequencing

The presence or absence of *mcr*-genes is determinable by PCR assays and whole-genome sequencing (WGS) as standard ^76^. Whilst WGS is able to characterize the mechanism of resistance and determine the molecular evolutionary trajectory of colistin-resistant isolates, PCR is only able to characterize resistance genes ^83, 84^ However, WGS technology is limited in settings that lack adequate resources and therefore PCR assays are widely adopted for detecting *mcr*-genes ^75, 85^.

Nijhuis *et al* (2016) were the first to design a real-time PCR assay for detecting *mcr*-1 from clinical isolates using self-designed primers and probes. The assay was validated on 26 *mcr*-1 positive *E. coli* isolates, where the presence of *mcr*-1 was detected in all 26 isolates ^86^. Additionally, the assay was evaluated on spiked stool samples and the efficiency of the PCR was 102.6% and the LOD was 3-30 cfu/reaction ^86^. However, *mcr*-1 genes were not detected in other colistin-resistant strains *i.e., Klebsiella, Enterobacter, Pseudomonas, Acinetobacter etc*.

A multiplex PCR (M-PCR) assay for the simultaneous detection of *mcr*-1 and carbapenem-resistant genes, *bla*_*KPC*_, *bla*_*NDM*_, *bla*_*IMP*_, *bla*_*OXA−48−like*_, was described ^87^. The assay was validated on reference strains including *E. coli* A434-59, which contains *mcr*-1 and *bla*_*NDM−1*_^87^. Evaluation of the M-PCR on 127 carbapenem-resistant, eight *mcr*-1-positive and 62 carbapenem-susceptible *Enterobacterales* found the assay to be 100% sensitive and specific ^87^.

Additionally, three studies designed M-PCR assays to detect *mcr*-1 to *mcr*-5 genes ^4, 85, 88^. The assay designed by Rebelo *et al* (2018) allowed for the simultaneous detection of *mcr* genes and their variants in bovine and porcine isolates ^85^. This study did not use internal amplification controls as they were incompatible with DreamTaq Green PCR Master Mix ^85^. The master mix contains DNA polymerase synthesized in *E. coli* and thus would produce amplicons if 16S rRNA primers are used ^85^.

However, Lescat *et al* (2018) designed a more rapid (<2 hours) M-PCR assay that was compatible with internal controls. Recently, Joussett *et al* (2019) designed and evaluated an M-PCR assay on 50 *E. coli*, 41 *K. pneumoniae* and 12 *Salmonella enterica* isolates (from which a total of 40 were MCR-producers), which was 100% accurate in detecting *mcr*-positive isolates. The assay was additionally performed on 82 *Aeromonas* spp. and 10 *Shewanella* spp. that were previously described as potential originators of *mcr*-3 and *mcr*-4, respectively ^4^. None of the *Aeromonas* spp. were *mcr*-positive, although two *Shewanella* spp., *S. bicestrii* JAB-1 strain and *S. woody* S539 with MICs of 0.25mg/L and <0.12 mg/L respectively, were *mcr*-4 positive ^4^. However, cloning *S. bicestrii* JAB-1 genes into *E. coli* TOP10 resulted in an *mcr-*4 positive outcome by the PCR assay with a colistin MIC of 4 mg/L ^4^.

Borowiak *et al* (2020) described the detection of *mcr*-1 to *mcr*-9 in colistin resistant *Salmonella enterica* isolates using an M-PCR (*mcr*-1 to *mcr*-5) designed by Rebello *et al* (2018) and a newly designed M-PCR assay (*mcr*-6 to *mcr*-9). The assay was performed on 407 colistin-resistant *S. enterica* isolates from animals, animal feed, food and the environment ^89^. Two hundred and fifty-four of the isolates had *mcr* genes. Moreover, the assay detected *mcr*-9 in isolates carrying *mcr*-1 ^89^. However, two separate frameshift mutations of *mcr*-9 were shown to have occurred in the respective isolates as demonstrated by WGS analysis; the mutations are believed to have contributed to non-functional MCR-9 proteins ^89^.

Two studies have described methods for broth enrichment of colistin-resistant *E. coli* followed by real-time PCR to detect *mcr*-genes ^90, 91^. Chalmers *et al* (2018) were the first to describe a SYBR Green-based real-time PCR method for *mcr*-1 and *mcr*-2 following enrichment with *E. coli* (EC) broth containing colistin (1µg/mL). All the porcine faecal and chicken caecal samples were screened by real-time PCR after 16h of culture in EC broth ^91^. However, none of the *mcr*-1 and *mcr*-2 genes were detected by PCR in any of the samples after 16h of enrichment ^91^. As well, the method described by Chandler *et al* (2020) for detecting *mcr*-1 included enrichment using EC broth containing colistin (1µg/mL) and vancomycin (8µg/mL). The method was evaluated on 100 feral swine faecal samples, which were inoculated with one of five different *mcr*-1 positive *E. coli* strains ^90^. The bacteria was inoculated at concentrations ranging between 0.1-9.99 CFU/g, 10-49.99 CFU/g, 50-99 CFU/g, 100-149 CFU/g and 200-2,200 CFU/g from which *mcr*-1 was detected with 32%, 72%, 88%, 95%, and 98% accuracy by real-time PCR, respectively ^90^.

Four SYBR Green-based real-time PCR assays have been developed for *mcr*-1, *mcr*-2, *mcr*-3, *mcr*-4, and *mcr*-5 detection ^92-95^. Bontron *et al* (2016) designed a SYBR Green-based real-time PCR assay for detection of *mcr*-1 from cultured bacteria and stools. The assay was validated on 20 *Enterobacterales,* where it was found to accurately detect the presence or absence of *mcr*-1 at a LOD of 10^2^ cultured bacteria ^93^. Furthermore, Dona *et al* (2017) described a SYBR Green real-time PCR assay to also detect *mcr*-1 from human faecal samples. However, in this study, 20µg of the stool samples were enriched overnight in 10mL LB broth containing 2µg/mL colistin and plated on four selective agar plates prior to DNA extraction ^95^. The real-time PCR accurately identified *mcr*-1 harbouring *E. coli* isolates with an LOD of 10^1^ and PCR efficiency of ca. 106% ^95^.

Li *et al* (2017) also designed a multiplex SYBR Green-based real-time PCR assay for *mcr*-1, *mcr*-2 and *mcr*-3 detection. The assay was validated on 25 isolates including *mcr*-1 positive and *mcr*-3 positive strains; the *mcr*-2 gene was synthesized in the study due to a lack of *mcr*-2-positive isolates ^94^. Although the *mcr* genes were detected with 100% accuracy with a LOD of 10^2^, *mcr*-2 was not validated on cultured bacteria ^94^. However, in this study all three *mcr* genes could not be simultaneously detected in one reaction unlike when the Taqman probe was used ^94^. A more recent study evaluated the SYBR Green-based real-time PCR method proposed by Li *et al* (2017) in detecting and quantifying *mcr*-1 to *mcr*-3 as well as newly designed assays for *mcr*-4 and *mcr*-5 ^92^. The optimized *mcr*-1 to *mcr*-5 PCR assays were validated on bacterial isolates ^92^. The study found that SYBR Green real-time PCR, followed by melting curve analysis, was more efficient in detecting and quantifying *mcr*-1 to *mcr*-5 genes in both bacterial isolates ^92^. The described assays detected all five *mcr* genes with a lower limit of 10^2^. Moreover, the assays enabled screening of five individual samples in a single reaction ^92^.

The parallel detection of *mcr*-1, *mcr*-2 and *mcr*-8 by real-time PCR using Taqman® probes has been described ^96-98^. Chabou *et al* (2016) designed two quantitative real-time PCR assays with TaqMan® probes for the rapid detection of *mcr*-1 gene. Primers and probes were designed to develop the two PCR assays, designated PE1 and PE2 ^96^. The assays were evaluated on 100 bacterial isolates (18 of which were colistin resistant) and 833 broiler faecal samples ^96^. The sensitivity and specificity of both assays were 100%, with a calibration curve that was linear from 10^1^ to 10^8^. However, the PE1 assay was recommended for initial screening of *mcr*-1 followed by PE2 assay for confirming the results ^96^.

Daniels *et al* (2017) developed a multiplexed real-time PCR with TaqMan® probes to detect *mcr*-1 and *mcr*-2. The assay was validated on 25 bacterial isolates, some of which were *mcr*-positive ^98^. The sensitivity and specificity of the assay was 100%, being able to detect *mcr*-1 and *mcr*-2 from dilutions containing 8.5 10^3^ and 7.7 10^3^cfu/mL respectively ^98^. A specific real-time PCR assay using TaqMan probes to identify *mcr*-8 was designed for the first time by Nabti *et al* (2020). The specificity and sensitivity of the assay were evaluated on 290 bacterial isolates from clinical samples and 250 metagenomic DNA from human stools ^97^. The PCR assay accurately detected *mcr*-8 from the one positive *K. pneumoniae* isolate with an overall efficiency of 92.64% and a limit of detection of 55 CFU/mL ^97^.

## 4. Conclusion

The rapid dissemination of colistin resistance, mediated by chromosomal mutations and *mcr*-genes, poses a threat to public and veterinary health as colistin is one of the last-line antibiotics. It is important to establish a simple, rapid, and cost-effective diagnostic method that will not discriminate against the mechanism of colistin resistance as well as take into consideration hetero-resistant isolates. Among the available diagnostic assays, the Rapid Resapolymyxin NP test is a promising initial screening method as it can be performed in-house, therefore making it relatively cheap; it is easy to perform and it is not limited to glucose-fermenting colistin-resistant Gram-negative bacteria. Other colorimetric screening methods such as ASAT, Rapid Polymyxin NP, Rapid Polymyxin *Acinetobacter,* and Rapid Polymyxin *Pseudomonas* are species-specific and cannot be used for general screening in high-capacity clinical laboratories.

Likewise, agar-based methods are cheap and can be used as initial screening tools in poorer settings, although most of the agar-based assays fail to detect isolates with hetero-resistance. They also have a longer TAT of 24 hours. Should there be a need to use agar-based assays, CHROMagar COL-*APSE* was designed to detect and differentiate all colistin-resistant isolates, although it might be relatively expensive than Superpolymyxin and the LBJMR medium, which can be performed in-house. However, the LBJMR medium was found to detect hetero-resistance better than Superpolymyxin. ChromID® Colistin R and Luria-Bertani (LB) media can only be used to screen for *Enterobacterales*.

MIC determiners could also be used for initial screening in highly resourced laboratories, although they relatively require a higher skill than the agar-based tests and the biochemical colorimetric tests. The non-automated MIC strips *i.e*., UMIC, MMS, and ComASP are cheaper than automated MIC determiners *i.e*., Microscan, Sensititre, and BD Phoenix. However, automated MIC determiners could be available in most well-resourced clinical laboratories as they are generally used for AST. Non-automated MIC strips are cheaper, require less skill, and recommendable for less-resourced laboratories.

A second screening can be performed to mainly detect *mcr*-production using chelator-based phenotypic assays, which are more suitable although most are subjected to >16h incubation. Moreover, the Lateral Flow assay that detects MCR-1/-2 production could be used for rapid detection. Molecular methods could be considered for the detection of *mcr*-genes in well-resourced clinical microbiology laboratories. Particularly, the LAMP assays could be used as they were found to be more sensitive than PCR methods. More so, LAMP requires less equipment and has a shorter TAT than PCR and WGS methods.

## Data Availability

All data are available in the manuscript

## Acknowledgements

none. We hereby regretfully report the death of our colleague Professor Nontombi Mbelle, who died during the submission of this article. This work is dedicated to her memory.

## Transparency declaration

Authors have no conflict of interest to declare.

## References

1. Vasoo, S. Susceptibility Testing for the Polymyxins: Two Steps Back, Three Steps Forward? Journal of clinical microbiology 55, 2573–2582 (2017).

2. Teo, J.W., Octavia, S., Cheng, J.W. & Lin, R.T., Vol. 90 67–69 (2018).

3. Li, B. et al. An enzyme-free homogenous electrochemical assay for sensitive detection of the plasmid-mediated colistin resistance gene mcr-1. Analytical and bioanalytical chemistry 410, 4885–4893 (2018).

4. Jousset, A.B. et al. Development and validation of a multiplex polymerase chain reaction assay for detection of the five families of plasmid-encoded colistin resistance. Int J Antimicrob Agents 53, 302–309 (2019).

5. Osei Sekyere, J., Sephofane, A.K. & Mbelle, N.M. Comparative Evaluation of CHROMagar COL-APSE, MicroScan Walkaway, ComASPColistin, and Colistin MAC Test in Detecting Colistin-resistant Gram-Negative Bacteria. medRxiv, 2019.2012.2017.19015156 (2020).

6. Liu, Y.-Y. et al. Emergence of plasmid-mediated colistin resistance mechanism MCR-1 in animals and human beings in China: a microbiological and molecular biological study. The Lancet infectious diseases 16, 161–168 (2016).

7. Dortet, L. et al. Optimization of the MALDIxin test for the rapid identification of colistin resistance in Klebsiella pneumoniae using MALDI-TOF MS. J Antimicrob Chemother 75, 110–116 (2020).

8. Bialvaei, A.Z. & Samadi Kafil, H. Colistin, mechanisms and prevalence of resistance. Current Medical Research and Opinion 31, 707–721 (2015).

9. Sun, J., Zhang, H., Liu, Y.-H. & Feng, Y. Towards Understanding MCR-like Colistin Resistance. Trends in Microbiology 26, 794–808 (2018).

10. Poirel, L., Jayol, A. & Nordmann, P. Polymyxins: Antibacterial Activity, Susceptibility Testing, and Resistance Mechanisms Encoded by Plasmids or Chromosomes. Clinical microbiology reviews 30, 557–596 (2017).

11. Nordmann, P., Jayol, A. & Poirel, L. Rapid Detection of Polymyxin Resistance in Enterobacteriaceae. Emerging infectious diseases 22, 1038–1043 (2016).

12. Osei Sekyere, J. Mcr colistin resistance gene: a systematic review of current diagnostics and detection methods. Microbiologyopen 8, e00682 (2019).

13. Rodriguez, C.H. et al. In-house rapid colorimetric method for detection of colistin resistance in Enterobacterales: A significant impact on resistance rates. Journal of chemotherapy (Florence, Italy) 31, 432–435 (2019).

14. Xavier, B.B. et al. Identification of a novel plasmid-mediated colistin-resistance gene, mcr-2, in Escherichia coli, Belgium, June 2016. Euro surveillance : bulletin Europeen sur les maladies transmissibles = European communicable disease bulletin 21 (2016).

15. AbuOun, M. et al. mcr-1 and mcr-2 variant genes identified in Moraxella species isolated from pigs in Great Britain from 2014 to 2015. The Journal of antimicrobial chemotherapy 72, 2745–2749 (2017).

16. Borowiak, M. et al. Identification of a novel transposon-associated phosphoethanolamine transferase gene, mcr- 5, conferring colistin resistance in d-tartrate fermenting Salmonella enterica subsp. enterica serovar Paratyphi B. The Journal of antimicrobial chemotherapy 72, 3317–3324 (2017).

17. Carattoli, A. et al. Novel plasmid-mediated colistin resistance mcr-4 gene in Salmonella and Escherichia coli, Italy 2013, Spain and Belgium, 2015 to 2016. Euro surveillance : bulletin Europeen sur les maladies transmissibles = European communicable disease bulletin 22 (2017).

18. Carroll, L.M. et al. Identification of Novel Mobilized Colistin Resistance Gene mcr-9 in a Multidrug-Resistant, Colistin-Susceptible Salmonella enterica Serotype Typhimurium Isolate. mBio 10 (2019).

19. Wang, X. et al. Emergence of a novel mobile colistin resistance gene, mcr-8, in NDM-producing Klebsiella pneumoniae. Emerging microbes & infections 7, 122 (2018).

20. Wenjuan, Y. et al. in mBio, Vol. 8 (2017).

21. Yong-Qiang, Y. et al. Novel plasmid-mediated colistin resistance gene mcr-7.1 in Klebsiella pneumoniae. Journal of Antimicrobial Chemotherapy (JAC) 73 (2018).

22. Furniss, R.C.D. et al. Detection of Colistin Resistance in Escherichia coli by Use of the MALDI Biotyper Sirius Mass Spectrometry System. Journal of clinical microbiology 57 (2019).

23. Bardet, L. & Rolain, J.-M. Development of New Tools to Detect Colistin-Resistance among Enterobacteriaceae Strains. Can J Infect Dis Med Microbiol 2018, 3095249–3095249 (2018).

24. Furniss, R.C.D. et al. Detection of Colistin Resistance in &lt;span class=&quot;named-content genus-species&quot; id=&quot;named-content-1&quot;&gt;Escherichia coli&lt;/span&gt; by Use of the MALDI Biotyper Sirius Mass Spectrometry System. Journal of Clinical Microbiology 57, e01427–01419 (2019).

25. Chew, K.L., La, M.V., Lin, R.T.P. & Teo, J.W.P. Colistin and Polymyxin B Susceptibility Testing for Carbapenem-Resistant and mcr-Positive Enterobacteriaceae: Comparison of Sensititre, MicroScan, Vitek 2, and Etest with Broth Microdilution. J Clin Microbiol 55, 2609–2616 (2017).

26. Jayol, A., Nordmann, P., Andre, C., Poirel, L. & Dubois, V. Evaluation of three broth microdilution systems to determine colistin susceptibility of Gram-negative bacilli. The Journal of antimicrobial chemotherapy 73, 1272–1278 (2018).

27. Simar, S., Sibley, D., Ashcraft, D. & Pankey, G. Colistin and Polymyxin B Minimal Inhibitory Concentrations Determined by Etest Found Unreliable for Gram-Negative Bacilli. The Ochsner journal 17, 239–242 (2017).

28. Haeili, M., Kafshdouz, M., Pishnian, Z., Feizabadi, M.M. & Martinez-Martinez, L. Comparison of susceptibility testing methods for determining the activity of colistin against Gram-negative bacilli of clinical origin. Journal of medical microbiology 68, 60–66 (2019).

29. Lutgring, J.D. et al. Evaluation of the MicroScan Colistin Well and Gradient Diffusion Strips for Colistin Susceptibility Testing in &lt;em&gt;Enterobacteriaceae&lt;/em&gt. Journal of Clinical Microbiology 57, e01866–01818 (2019).

30. Matuschek, E., Åhman, J., Webster, C. & Kahlmeter, G. Antimicrobial susceptibility testing of colistin–evaluation of seven commercial MIC products against standard broth microdilution for Escherichia coli, Klebsiella pneumoniae, Pseudomonas aeruginosa, and Acinetobacter spp. Clinical Microbiology and Infection 24, 865–870 (2018).

31. Mitton, B., Kingsburgh, C., Kock, M., Mbelle, N. & Strydom, K. Evaluation of an In-House Colistin NP Test for Use in Resource-Limited Settings. Journal of clinical microbiology 57, e00501–00519 (2019).

32. Jayol, A. et al. Hafnia, an enterobacterial genus naturally resistant to colistin revealed by three susceptibility testing methods. The Journal of antimicrobial chemotherapy 72, 2507–2511 (2017).

33. Bardet, L., Okdah, L., Le Page, S., Baron, S.A. & Rolain, J.M. Comparative evaluation of the UMIC Colistine kit to assess MIC of colistin of gram-negative rods. BMC Microbiol 19, 60 (2019).

34. Carretto, E. et al. Clinical Validation of SensiTest Colistin, a Broth Microdilution-Based Method To Evaluate Colistin MICs. Journal of Clinical Microbiology 56, e01523–01517 (2018).

35. Hong, J.S. et al. Evaluation of the BD Phoenix M50 Automated Microbiology System for Antimicrobial Susceptibility Testing with Clinical Isolates in Korea. Microb Drug Resist 25, 1142–1148 (2019).

36. Malli, E. et al. Evaluation of rapid polymyxin NP test to detect colistin-resistant Klebsiella pneumoniae isolated in a tertiary Greek hospital. Journal of microbiological methods 153, 35–39 (2018).

37. Jayol, A., Nordmann, P., Lehours, P., Poirel, L. & Dubois, V. Comparison of methods for detection of plasmidmediated and chromosomally encoded colistin resistance in Enterobacteriaceae. Clinical Microbiology and Infection 24, 175–179 (2018).

38. Rodriguez, C.H. et al. Discrepancies in susceptibility testing to colistin in Acinetobacter baumannii: The influence of slow growth and heteroresistance. International journal of antimicrobial agents 54, 587–591 (2019).

39. Poirel, L. et al. Rapid Polymyxin NP test for the detection of polymyxin resistance mediated by the mcr-1/mcr-2 genes. Diagnostic microbiology and infectious disease 90, 7–10 (2018).

40. Yainoy, S. et al. Evaluation of the Rapid Polymyxin NP test for detection of colistin susceptibility in Enterobacteriaceae isolated from Thai patients. Diagn Microbiol Infect Dis 92, 102–106 (2018).

41. Jayol, A. et al. Evaluation of the Rapid Polymyxin NP test and its industrial version for the detection of polymyxinresistant Enterobacteriaceae. Diagn Microbiol Infect Dis 92, 90–94 (2018).

42. Jayol, A., Dubois, V., Poirel, L. & Nordmann, P. Rapid Detection of Polymyxin-Resistant Enterobacteriaceae from Blood Cultures. Journal of clinical microbiology 54, 2273–2277 (2016).

43. Malli, E., Papagiannitsis, C.C., Xitsas, S., Tsilipounidaki, K. & Petinaki, E. Implementation of the Rapid Polymyxin™ NP test directly to positive blood cultures bottles. Diagnostic Microbiology and Infectious Disease 95, 114889 (2019).

44. Przybysz, S.M., Correa-Martinez, C., Kock, R., Becker, K. & Schaumburg, F. SuperPolymyxin Medium for the Screening of Colistin-Resistant Gram-Negative Bacteria in Stool Samples. Frontiers in microbiology 9, 2809 (2018).

45. Dalmolin, T.V. et al. Detection of Enterobacterales resistant to polymyxins using Rapid Polymyxins NP test. Braz J Microbiol 50, 425–428 (2019).

46. Malli, E. et al. Implementation of the Rapid Polymyxin Acinetobacter Test to Detect Colistin-Resistant Acinetobacter baumannii. Microbial drug resistance (Larchmont, N.Y.) (2019).

47. Lescat, M., Poirel, L., Jayol, A. & Nordmann, P. Performances of the Rapid Polymyxin Acinetobacter and Pseudomonas Tests for Colistin Susceptibility Testing. Microb Drug Resist 25, 520–523 (2019).

48. Sadek, M., Tinguely, C., Poirel, L. & Nordmann, P. Rapid Polymyxin/Pseudomonas NP test for rapid detection of polymyxin susceptibility/resistance in Pseudomonas aeruginosa. Eur J Clin Microbiol Infect Dis 39, 1657–1662 (2020).

49. Lescat, M., Poirel, L., Tinguely, C. & Nordmann, P. A Resazurin Reduction-Based Assay for Rapid Detection of Polymyxin Resistance in Acinetobacter baumannii and Pseudomonas aeruginosa. J Clin Microbiol 57 (2019).

50. Jia, H. et al. Evaluation of resazurin-based assay for rapid detection of polymyxin-resistant gram-negative bacteria. BMC Microbiol 20, 7 (2020).

51. Germ, J. et al. Evaluation of resazurin-based rapid test to detect colistin resistance in Acinetobacter baumannii isolates. Eur J Clin Microbiol Infect Dis 38, 2159–2162 (2019).

52. Esposito, F. et al. Detection of Colistin-Resistant MCR-1-Positive Escherichia coli by Use of Assays Based on Inhibition by EDTA and Zeta Potential. J Clin Microbiol 55, 3454–3465 (2017).

53. Yauri Condor, K., Gonzales Escalante, E., Di Conza, J. & Gutkind, G. Detection of plasmid-mediated colistin resistance by colistin pre-diffusion and inhibition with EDTA test (CPD-E) in Enterobactereaceae. Journal of Microbiological Methods 167 (2019).

54. Clément, M. et al. The EDTA-based disk-combination tests are unreliable for the detection of MCR-mediated colistin-resistance in Enterobacteriaceae. Journal of Microbiological Methods 153, 31–34 (2018).

55. Budel, T. et al. Evaluation of EDTA- and DPA-Based Microdilution Phenotypic Tests for the Detection of MCR-Mediated Colistin Resistance in Enterobacteriaceae. Microb Drug Resist 25, 494–500 (2019).

56. Simner, P.J. et al. Two-Site Evaluation of the Colistin Broth Disk Elution Test To Determine Colistin &lt;em&gt;In Vitro&lt;/em&gt; Activity against Gram-Negative Bacilli. Journal of Clinical Microbiology 57, e01163–01118 (2019).

57. Dalmolin, T.V. et al. Elution methods to evaluate colistin susceptibility of Gram-negative rods. Diagnostic Microbiology and Infectious Disease 96, 114910 (2020).

58. Fenwick, A.J. et al. Evaluation of the NG-Test MCR-1 Lateral Flow Assay and EDTA-Colistin Broth Disk Elution Methods To Detect Plasmid-Mediated Colistin Resistance Among Gram-Negative Bacterial Isolates. Journal of Clinical Microbiology, JCM.01823-01819 (2020).

59. Bell, D.T. et al. A Novel Phenotypic Method To Screen for Plasmid-Mediated Colistin Resistance among &lt;em&gt;Enterobacteriales&lt;/em&gt. Journal of Clinical Microbiology 57, e00040–00019 (2019).

60. Coppi, M. et al. A simple phenotypic method for screening of MCR-1-mediated colistin resistance. Clinical microbiology and infection : the official publication of the European Society of Clinical Microbiology and Infectious Diseases 24, 1–201 (2018).

61. Jouy, E. et al. Improvement in routine detection of colistin resistance in E. coli isolated in veterinary diagnostic laboratories. Journal of microbiological methods 132, 125–127 (2017).

62. Volland, H. et al. Development and Multicentric Validation of a Lateral Flow Immunoassay for Rapid Detection of MCR-1-Producing Enterobacteriaceae. Journal of clinical microbiology 57 (2019).

63. Abdul Momin, M.H.F. et al. CHROMagar COL-APSE: a selective bacterial culture medium for the isolation and differentiation of colistin-resistant Gram-negative pathogens. J Med Microbiol 66, 1554–1561 (2017).

64. Girlich, D., Naas, T. & Dortet, L. Comparison of the Superpolymyxin and ChromID Colistin R Screening Media for the Detection of Colistin-Resistant Enterobacteriaceae from Spiked Rectal Swabs. Antimicrob Agents Chemother 63 (2019).

65. Nordmann, P., Jayol, A. & Poirel, L. A Universal Culture Medium for Screening Polymyxin-Resistant Gram-Negative Isolates. Journal of clinical microbiology 54, 1395–1399 (2016).

66. Bardet, L., Page, S.L., Leangapichart, T. & Rolain, J.-M. LBJMR medium: a new polyvalent culture medium for isolating and selecting vancomycin and colistin-resistant bacteria. BMC microbiology 17, 220 (2017).

67. Jayol, A., Poirel, L., Andre, C., Dubois, V. & Nordmann, P. Detection of colistin-resistant Gram-negative rods by using the SuperPolymyxin medium. Diagn Microbiol Infect Dis 92, 95–101 (2018).

68. van Hout, D. et al. The added value of the selective SuperPolymyxin(tm) medium in detecting rectal carriage of Gram-negative bacteria with acquired colistin resistance in intensive care unit patients receiving selective digestive decontamination. Eur J Clin Microbiol Infect Dis 39, 265–271 (2020).

69. Germ, J. & Seme, K.a. Evaluation of a novel epidemiological screening approach for detection of colistin resistant human Enterobacteriaceae isolates using a selective SuperPolymyxin medium. Journal of microbiological methods 160, 117–123 (2019).

70. Thiry, D. et al. Assessment of two selective agar media to isolate colistin-resistant bovine Escherichia coli: Correlation with minimal inhibitory concentration and presence of mcr genes. Journal of Microbiological Methods 159, 174–178 (2019).

71. García-Fernández, S. et al. Performance of CHROMID® Colistin R agar, a new chromogenic medium for screening of colistin-resistant Enterobacterales. Diagnostic Microbiology and Infectious Disease 93, 1–4 (2019).

72. Turbett, S.E. et al. Evaluation of a Screening Method for the Detection of Colistin-Resistant Enterobacteriaceae in Stool. Open forum infectious diseases 6, ofz211 (2019).

73. Girlich, D. et al. Evaluation of the Amplidiag CarbaR+MCR Kit for Accurate Detection of Carbapenemase-Producing and Colistin-Resistant Bacteria. Journal of Clinical Microbiology 57, e01800–01818 (2019).

74. Meunier, D., Woodford, N. & Hopkins, K.L. Evaluation of the AusDiagnostics MT CRE EU assay for the detection of carbapenemase genes and transferable colistin resistance determinants mcr-1/-2 in MDR Gram-negative bacteria. The Journal of antimicrobial chemotherapy 73, 3355–3358 (2018).

75. Zhong, L.-L., Zhou, Q., Tan, C.-Y. & Roberts, A.P.a. Multiplex loop-mediated isothermal amplification (multi-LAMP) assay for rapid detection of mcr-1 to mcr-5 in colistin-resistant bacteria. Infect Drug Resist 12, 1877–1887 (2019).

76. Imirzalioglu, C. et al., Vol. 61 (2017).

77. Zou, D. et al. Sensitive and Rapid Detection of the Plasmid-Encoded Colistin-Resistance Gene mcr-1 in Enterobacteriaceae Isolates by Loop-Mediated Isothermal Amplification. Frontiers in microbiology 8, 2356 (2017).

78. Giordano, C. & Barnini, S. Rapid detection of colistin-resistant Klebsiella pneumoniae using MALDI-TOF MS peak-based assay. J Microbiol Methods 155, 27–33 (2018).

79. Dortet, L. et al. Rapid detection of colistin resistance in Acinetobacter baumannii using MALDI-TOF-based lipidomics on intact bacteria. Scientific reports 8, 16910 (2018).

80. Bernasconi, O.J. et al., Vol. 55 3138–3141 (2017).

81. Gong, L. et al. Multiple Cross Displacement Amplification Coupled With Gold Nanoparticles-Based Lateral Flow Biosensor for Detection of the Mobilized Colistin Resistance Gene mcr-1. Frontiers in cellular and infection microbiology 9, 226 (2019).

82. Xu, J., Wang, X., Yang, L., Kan, B. & Lu, X. Rapid detection of mcr-1 by recombinase polymerase amplification. Journal of medical microbiology 67, 1682–1688 (2018).

83. Peter, S. et al. Whole-genome sequencing enabling the detection of a colistin-resistant hypermutating Citrobacter werkmanii strain harbouring a novel metallo-beta-lactamase VIM-48. International journal of antimicrobial agents 51, 867–874 (2018).

84. Hua, X. et al. Colistin Resistance in Acinetobacter baumannii MDR-ZJ06 Revealed by a Multiomics Approach. Frontiers in cellular and infection microbiology 7, 45 (2017).

85. Rebelo, A.R. et al. Multiplex PCR for detection of plasmid-mediated colistin resistance determinants, mcr-1, mcr-2, mcr-3, mcr-4 and mcr-5 for surveillance purposes. Euro surveillance : bulletin Europeen sur les maladies transmissibles = European communicable disease bulletin 23 (2018).

86. Nijhuis, R. et al. Detection of the plasmid-mediated colistin-resistance gene mcr-1 in clinical isolates and stool specimens obtained from hospitalized patients using a newly developed real-time PCR assay. Journal of Antimicrobial Chemotherapy 71, 2344–2346 (2016).

87. Hatrongjit, R., Kerdsin, A., Akeda, Y. & Hamada, S. Detection of plasmid-mediated colistin-resistant and carbapenem-resistant genes by multiplex PCR. MethodsX 5, 532–536 (2018).

88. Lescat, M., Poirel, L. & Nordmann, P. Rapid multiplex polymerase chain reaction for detection of mcr-1 to mcr-5 genes. Diagnostic Microbiology and Infectious Disease 92, 267–269 (2018).

89. Borowiak, M. et al. Development of a Novel mcr-6 to mcr-9 Multiplex PCR and Assessment of mcr-1 to mcr-9 Occurrence in Colistin-Resistant Salmonella enterica Isolates From Environment, Feed, Animals and Food (2011– 2018) in Germany. Frontiers in Microbiology 11 (2020).

90. Chandler, J.C. et al. Validation of a screening method for the detection of colistin-resistant E. coli containing mcr-1 in feral swine feces. Journal of microbiological methods 172, 105892 (2020).

91. Chalmers, G. et al. A method to detect Escherichia coli carrying the colistin-resistance genes mcr-1 and mcr-2 using a single real-time polymerase chain reaction and its application to chicken cecal and porcine fecal samples. Canadian journal of veterinary research = Revue canadienne de recherche veterinaire 82, 312–315 (2018).

92. Tolosi, R. et al. Rapid detection and quantification of plasmid-mediated colistin resistance genes (mcr-1 to mcr-5) by real-time PCR in bacterial and environmental samples. Journal of Applied Microbiology n/a (2020).

93. Bontron, S.v., Poirel, L. & Nordmann, P. Real-time PCR for detection of plasmid-mediated polymyxin resistance (mcr-1) from cultured bacteria and stools. Journal of Antimicrobial Chemotherapy (JAC) 71 (2016).

94. Li, J. et al. A Multiplex SYBR Green Real-Time PCR Assay for the Detection of Three Colistin Resistance Genes from Cultured Bacteria, Feces, and Environment Samples. Frontiers in microbiology 8, 2078 (2017).

95. Dona, V., Bernasconi, O.J., Kasraian, S., Tinguely, R. & Endimiani, A. A SYBR(®) Green-based real-time PCR method for improved detection of mcr-1-mediated colistin resistance in human stool samples. J Glob Antimicrob Resist 9, 57–60 (2017).

96. Chabou, S., Leangapichart, T. & Okdah, L.a. Real-time quantitative PCR assay with Taqm®(R)) probe for rapid detection of MCR-1 plasmid-mediated colistin resistance. New microbes and new infections 13, 71–74 (2016).

97. Nabti, L.Z. et al. Development of real-time PCR assay allowed describing the first clinical Klebsiella pneumoniae isolate harboring plasmid-mediated colistin resistance mcr-8 gene in Algeria. Journal of global antimicrobial resistance 20, 266–271 (2020).

98. Daniels, J.B. et al. Development and Validation of a Clinical Laboratory Improvement Amendments-Compliant Multiplex Real-Time PCR Assay for Detection of mcr Genes. Microb Drug Resist 25, 991–996 (2019).

